# Interaction between myelodysplasia-related gene mutations and ontogeny in acute myeloid leukemia: an appraisal of the new WHO and IC classifications and ELN risk stratification

**DOI:** 10.1101/2022.12.26.22283876

**Authors:** Joseph GW. McCarter, David Nemirovsky, Christopher A. Famulare, Noushin Farnoud, Abhinita S. Mohanty, Zoe S. Stone-Molloy, Jordan Chervin, Brian J. Ball, Zachary D. Epstein-Peterson, Maria E. Arcila, Aaron J. Stonestrom, Andrew Dunbar, Sheng F. Cai, Jacob L. Glass, Mark B. Geyer, Raajit K. Rampal, Ellin Berman, Omar I. Abdel-Wahab, Eytan M. Stein, Martin S. Tallman, Ross L. Levine, Aaron D. Goldberg, Elli Papaemmanuil, Yanming Zhang, Mikhail Roshal, Andriy Derkach, Wenbin Xiao

**Affiliations:** Computational Oncology Service, Department of Epidemiology & Biostatistics; MSK Kids; Center for Hematologic Malignancies; Biostatistics Service, Department of Epidemiology & Biostatistics; Diagnostic Molecular Laboratory, Department of Pathology and Laboratory Medicine; Leukemia Service, Department of Medicine; Molecular Cancer Medicine Service, Human Oncogenesis & Pathogenesis Program; Cytogenetics Laboratory, Department of Pathology and Laboratory Medicine; Hematopathology Service, Department of Pathology and Laboratory Medicine Memorial Sloan Kettering Cancer Center, New York, NY, USA

**Author notes:** Correspondence: Wenbin Xiao. Z.S.S. is at the Jacobs School of Medicine and Biomedical Sciences at the University at Buffalo, Buffalo, NY. J.C. is at Weill Cornell Medicine, New York City, NY. B.B. is at City of Hope National Medical Center, Duarte, CA. M.S.T. is at Northwestern University, Chicago, IL.

**Keywords:** acute myeloid leukemia, ontogeny, myelodysplasia-related, *de novo*, therapy-related, mutations, genomics, risk stratification, classification, outcome

## Abstract

Accurate classification and risk stratification is critical for clinical decision making in AML patients. In the newly proposed World Health Organization (WHO) and International Consensus classifications (ICC) of hematolymphoid neoplasms, the presence of myelodysplasia-related (MR) gene mutations is included as one of the diagnostic criteria of AML, myelodysplasia-related (AML-MR), largely based on the assumption that these mutations are specific for AML with an antecedent myelodysplastic syndrome. ICC also prioritizes MR gene mutations over ontogeny (as defined by clinical history). Furthermore, European LeukemiaNet (ELN) 2022 stratifies these MR gene mutations to the adverse-risk group. By thoroughly annotating a cohort of 344 newly diagnosed AML patients treated at Memorial Sloan Kettering Cancer Center (MSKCC), we show that ontogeny assignment based on database registry lacks accuracy. MR gene mutations are frequently seen in *de novo* AML. Among MR gene mutations, only *EZH2* and *SF3B1* were associated with an inferior outcome in a univariate analysis. In a multivariate analysis, AML ontogeny had independent prognostic values even after adjusting for age, treatment, allo-transplant and genomic classes or ELN risks. Ontogeny also stratified the outcome of AML with MR gene mutations. Finally, *de novo* AML with MR gene mutations did not show an adverse outcome. In summary, our study emphasizes the importance of accurate ontogeny designation in clinical studies, demonstrates the independent prognostic value of AML ontogeny and questions the current classification and risk stratification of AML with MR gene mutations.

**Key points:**

- Both ontogeny and genomics show independent prognostic values in AML.
- The newly proposed myelodysplasia-related gene mutations are neither specific to AML-MRC^WHO2016^ nor predictive for adverse outcomes.
- Ontogeny stratifies the outcome of AML with myelodysplasia-related gene mutations.

## Introduction

Acute myeloid leukemia (AML) is a heterogeneous group of clinically aggressive hematologic malignancies characterized by maturation arrest and accumulation of myeloid blasts^1,2^. AML classification is a prerequisite for appropriate disease management and has been an evolving process. The French-American-British (FAB) classification, arguably the first widely adapted classification scheme was largely based on cytomorphology and cytochemical characteristics, which, despite limitations, informed the formulation of the modern AML classification^3-5^. With a deeper understanding of the underlying molecular pathogenesis, it has become clear that an integrated algorithm including clinical history, morphology, immunophenotype and genetics provides better disease delineation which can further guide clinical decision making^6^. This culminated in the WHO 2016 classification of hematolymphoid neoplasms (WHO^2016^) that classifies AML into 4 major subtypes: AML with recurrent genetic abnormalities (AML-RGA), AML with myelodysplasia-related changes (AML-MRC^WHO2016^), therapy-related AML (t-AML) and AML, not otherwise specified (AML, NOS)^4^. Prior cytotoxic therapy for a non-myeloid disorder is a prerequisite for the diagnosis of t-AML. Diagnostic criteria for AML-MRC^WHO2016^ are: 1) history of myelodysplastic syndrome (MDS) or MDS/myeloproliferative neoplasm (MDS/MPN); or 2) the presence of MDS-defining cytogenetic abnormalities; or 3) morphologic dysplasia in greater than 50% of cells of at least 2 lineages. The evaluation of dysplasia appears subjective with a poor interobserver agreement; therefore, its value as a criterion of AML-MRC^WHO2016^ has been questioned^7-11^. Notably, AML ontogeny, as defined by clinical history outweighed genomics in WHO^2016^ classification.

The genomic landscape of AML has been increasingly delineated in the last decade^12-15^. Genomic classification of AML has thus been proposed based on the presence of mutations/fusions adding prognostic value to risk stratification^15,16^. Besides the well-recognized AML-RGA as defined by WHO^2016^, two large genomic clusters of patients with either *TP53* or chromatin-spliceosome mutations are also identified^15^, presumably corresponding to t-AML and AML-MRC^WHO2016^, respectively. Furthermore, a study based on a multi-center clinical trial demonstrated that mutations in 8 genes (i.e. *ASXL1, BCOR, EZH2, STAG2, SF3B1, SRSF2, ZRSR2, U2AF1*) within the chromatin-spliceosome cluster appear 95% specific to AML-MRC^WHO2016^ in comparison to *de novo* AML^17^. Based on these new data, both WHO^2022^ and ICC^2022^ include a new AML subtype classifying AMLs with the presence of any of these 8 gene mutations as AML with myelodysplasia-related gene mutations (MR genes)^18,19^. ICC also adds *RUNX1* mutations to the list (*MR/RUNX1*)^19^. Moreover, while WHO still retains ontogeny in the diagnostic hierarchy^18^, genomic features override ontogeny in ICC which means that ICC abandons t-AML and AML-MRC^WHO2016^ as diagnostic entities^19^. Concurrently, a few studies have shown inferior outcomes in AML patients carrying MR gene mutations ^16,20-23^. Therefore, these mutations have been universally added to the adverse-risk group by the ELN2022^2^. Despite of these new classifications/risk stratification, there is a lack of data demonstrating the relative importance of ontogeny versus genomics in AML prognosis. Thorough and comprehensive review of patient history is essential for an accurate ontogeny assignment, which unfortunately has been challenging in many published studies to date. Second, whether all the assigned MR gene mutations are specific to AML-MRC^WHO2016^ and are *bona fide* adverse risk factors as designated by ELN2022 awaits independent validation.

In this study, we manually annotated a cohort of 344 newly diagnosed AML patients treated at MSKCC and correlated ontogeny and genomics with outcomes. We show that: 1) ontogeny assignment from the database registry lacks sensitivity and specificity; 2) both ontogeny and genomics provide independent prognostic values in AML; 3) the newly proposed MR gene mutations are enriched in but not specific to AML-MRC^WHO2016^, and do not necessarily correlate with adverse outcomes; 4) *de novo* AML with MR gene mutations shows an outcome that falls in between favorable- and intermediate-risk groups.

## Methods

### Patient cohort

Patients with a newly diagnosed AML from January 2014 to December 2019 at MSKCC were retrospectively enrolled into this study. Any patients who lacked genomic data or follow ups were excluded. AML diagnosis was confirmed independently by two hematopathologists (M.R. and W.X.). Therapy-related AML (t-AML) was defined according to WHO^2016^ criteria as a late complication of cytotoxic chemotherapy and/or radiation therapy administered for a prior non-myeloid neoplastic or non-neoplastic disorder including alkylating agents, platinum derivatives, topoisomerase II inhibitors, anti-metabolites, anti-tubulin agents, external beam radiotherapy to active marrow sites, and therapeutic systemic radioisotopes^4^. AML-MRC^WHO2016^ was further divided into MR-Hx and MR-CG: AML was classified as MR-Hx when developed at least 3 months after the histologic documentation of antecedent MDS or MDS/MPN and not qualifying as t-AML. In the absence of the abovementioned history, AML with MDS-defining cytogenetic abnormalities was classified as MR-CG. Morphologic dysplasia was not included as one of the criteria for AML-MRC^WHO2016^ in this study. It should be noted that the criteria of t-AML, MR-Hx and MR-CG are nearly identical between WHO^2008^ and WHO^2016^. The remaining cases were classified as *de novo* AML. Therefore, AML ontogeny was assigned as t-AML, AML-MRC^WHO2016^ (specifically MR-Hx vs. MR-CG) and *de novo* AML. This study was approved by the Institutional Review Board at MSKCC.

### Chromosome and FISH analysis

Conventional chromosome analysis was performed on fresh bone marrow aspirate and/or peripheral blood specimens following standard protocol. At least 20 metaphase cells were analyzed, and karyotype was described according to the international system of human chromosome nomenclature (ISCN, 2016). FISH analysis was performed on bone marrow or peripheral blood pellets following standard protocols. Various commercial FISH probes, specific for myeloid neoplasia, such as for deletion or loss of chromosomes 5, 7, 17/*TP53*, gain of chromosome 8, deletion of 20q, and for *MLL/KMT2A* (11q23) translocations, *EVI1* (3q26.2), and other translocations, were used as appropriate. At least 300 cells were analyzed in each sample, and results were described according to ISCN, 2016. FISH results were correlated with chromosome analysis findings, when feasible also on metaphase cells for a precise interpretation of chromosome abnormalities. All cytogenetic karyotypes were manually re-annotated by a cytogeneticist (Y.Z.) based on numerical and structural chromosomal abnormalities, such as gain, loss, deletion, addition, duplication, balanced or unbalanced translocations, inversion, and derivative chromosomes, as well as marker chromosome, ring chromosome to define complex karyotypes (CK), i.e., three or more clonal chromosome abnormalities, clonal heterogeneity, and monosomal karyotype (MK).

### Mutation profiling

Next generation sequencing (NGS) studies were performed on submitted bone marrow or peripheral blood cells at diagnosis using one of the three clinically validated targeted panels: Raindance Thunderstorm (RDTS, 28-gene panel), Raindance Thunderbolt (RDTB, 49-gene panel) or IMPACT-heme (400-gene panel) (**supplemental Table 1**) ^24^. The detection of FLT3 mutations were supplemented by PCR tests. There were 23 patients who had NGS studies not at diagnosis but only performed on refractory/persistent disease (>20% blasts) within 4 months after the initiation of treatment. Patients who only had NGS data at relapse were excluded. Candidate mutations were annotated with VAGrENT (v. 3.3.0) (https://github.com/cancerit/VAGrENT) and Ensemble (v. 91) - VEP (v. 92) (https://github.com/Ensembl/ensembl-vep). They were compared to the COSMIC (v. 81)^25^, OncoKB^26^ and Genome Aggregation Database (gnomAD) ^27^ databases along with recurrence in a panel of normal samples to provide further information about the prevalence of each mutation in human cancer and normal populations. This information was used for the manual curation of each variant in order to classify it as pathogenic, likely pathogenic, or a variant of uncertain significance. Pathogenic and likely pathogenic mutations were retained for analysis and visualization (**supplemental Table 2**).

### Genomic class

Genomic class assignment was based on the WHO^2022^ and IC^2022^ classifications^18,19^. The patients were classified in an hierarchical order as well-defined AML entities with fusions including t(15;17), t(8;21), inv(16), t(6;9), *MLL* rearrangements, and *EVI1* rearrangements, followed by *NPM1, CEBPA bZIP, TP53*, and lastly *MR/RUNX1* mutations. Disease-defining fusions override disease-defining mutations when co-occurring. There were no patients with *NPM1* and *TP53* co-mutations. One patient with a *CEBPA bZIP* mutation and a subclonal *TP53* mutation was classified as AML with *CEBPA bZIP* mutation. AML with *MR/RUNX1* mutations is defined as the presence of any *MR/RUNX1* mutations but no disease defining-fusions or mutations (i.e. *NPM1, CEBPA bZIP* or *TP53*).

### Statistics

Descriptive statistics, including median and interquartile range [IQR] for continuous variables, and percentages for categorical variables, are provided. Fisher’s exact test or χ2 test was used to evaluate the association between two categorical variables. Wilcoxon rank-sum test or Kruskal-Wallis test was used to assess the difference in a continuous variable between/among patient groups. Univariable logistic regression was performed to assess the association between mutations profiles and AML ontogeny (MR-Hx vs. *de novo* AML, MR-Hx vs. t-AML and t-AML vs. *de novo* AML). Overall survival (OS) was calculated from the start of systematic treatment for the AML or the date of diagnosis for patients under supportive care until death or the date of the last follow-up. Left truncation was used to account for molecular testing performed after the start of first line chemotherapy in a small group of patients (n=23). Kaplan-Meier method was used to estimate OS and log-rank test to evaluated differences between groups. Univariable Cox proportional hazard regression to evaluate OS associated with disease-related determinants such as AML ontogeny and mutation profiles. We also evaluated AML ontogeny related risk in multivariable Cox-regression models adjusting for age (modeled by cubic spline), initial treatment at diagnosis, allogenic hematopoietic stem/progenitor cell (allo-HSCT) transplant (modeled as time-dependent variable), presence of *TP53* mutation, ELN2022 risk, cytogenetics, genomics, or sequencing platform. In sensitivity analysis, we stratified on ELN2022 risk, cytogenetics, genomics, or sequencing platform while adjusting for the same set of covariates to confirm association between AML ontology and OS. All analyses and graphics were produced using R version 4.0.3.

## Results

### Re-classification of AML ontogeny

The entire study cohort comprised 344 patients newly diagnosed as AML (male to female ratio: 1.4; median age: 66.7 years, **supplemental Table 3**). Based on the information extracted from the database registry, the ontogeny was assigned as follows: t-AML (n=48, 14.0%), and AML-MRC^WHO2016^ (n=139, 40.4%), and *de novo* AML (n=157, 45.6%). In order to accurately classify AML based on ontogeny and WHO^2016^ criteria, a multidisciplinary team curated each patient via a thorough chart review including the history of antecedent hematologic diseases, cancers, chemoradiation therapy, laboratory tests, and cytogenetic reports. Curated data was independently validated by two board-certified hematopathologists (M.R. and W.X.). Karyotyping and FISH results were re-annotated by a board-certified cytogeneticist (Y.Z.). Any discrepancies were resolved by group consensus.

To this end, the entire cohort was re-classified based on the thorough review into t-AML (92/344, 26.7%), MR-Hx (107/344, 31.1%), MR-CG (29/344, 8.4%), and *de novo* AML (116/344, 33.7%) (see Methods for definitions, **Table 1**). Although the initial classification of t-AML was highly specific, the sensitivity was only 52.2%. Half of t-AMLs were initially misclassified as AML-MRC^WHO2016^ or *de novo* AML due to an inadequate history review. The initial classification of AML-MRC^WHO2016^ was moderately specific (79.3%) and sensitive (70.6%). Importantly, the initial designation of *de novo* AML had a specificity of only 78.1% with nearly a third of these cases being t-AML or AML-MRC^WHO2016^. These findings demonstrate the importance of thorough cohort annotation to render an accurate assignment of AML ontogeny.

**Table 1.**
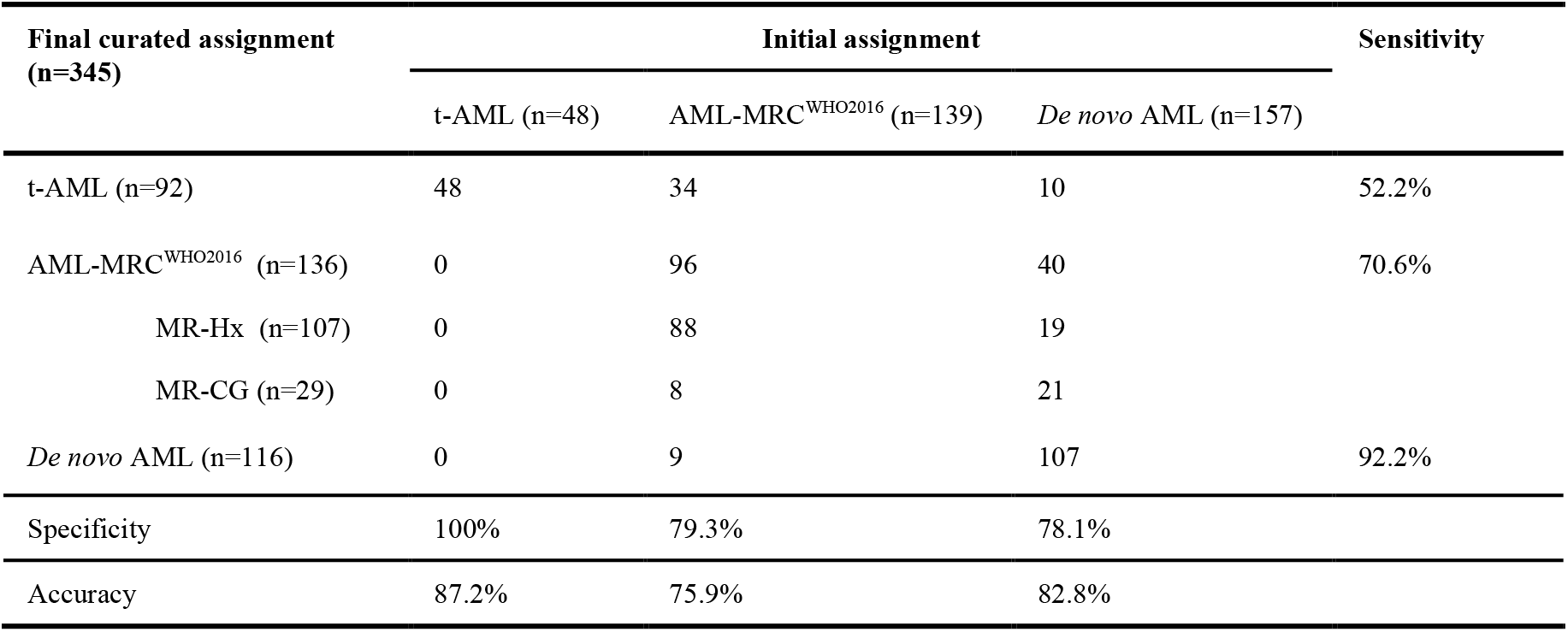
Reclassification of AML ontogeny.

| Final curated assignment<br>(n=345) | Initial assignment |  |  | Sensitivity |
| --- | --- | --- | --- | --- |
|  | t-AML (n=48) | AML-MRC<sup>WHO2016</sup> (n=139) | *De novo* AML (n=157) |  |
| t-AML (n=92) | 48 | 34 | 10 | 52.2% |
| AML-MRC<sup>WHO2016</sup> (n=136) | 0 | 96 | 40 | 70.6% |
| MR-Hx (n=107) | 0 | 88 | 19 |  |
| MR-CG (n=29) | 0 | 8 | 21 |  |
| *De novo* AML (n=116) | 0 | 9 | 107 | 92.2% |
| Specificity | 100% | 79.3% | 78.1% |  |
| Accuracy | 87.2% | 75.9% | 82.8% |  |

### Prediction of AML Ontogeny by mutations

The clinical characteristics of AML subtypes were shown in **Table 2**. Patients with *de novo* AML were younger than those with t-AML or MR-Hx (median age: 60 years vs 70 or 69 years, respectively, p<0.001). There were roughly two-fold more patients with *de novo* AML receiving induction chemotherapy and allo-transplant than those with t-AML and MR-Hx (**Table 2**). The median interval between MDS or MDS/MPN and MR-Hx was 15 months (IQR: 6-26 months). The median latency of t-AML from the prior chemoradiation therapy was 71 months (IQR: 29-106 months). A small proportion of the patients with *de novo* AML (8/116, 6.9%), MR-Hx (13/107, 12%) and MR-CG (4/29, 14%) also had a history of solid cancers mostly at early stages and treated with surgical resection solely for which none of whom received chemoradiation therapy.

**Table 2.**
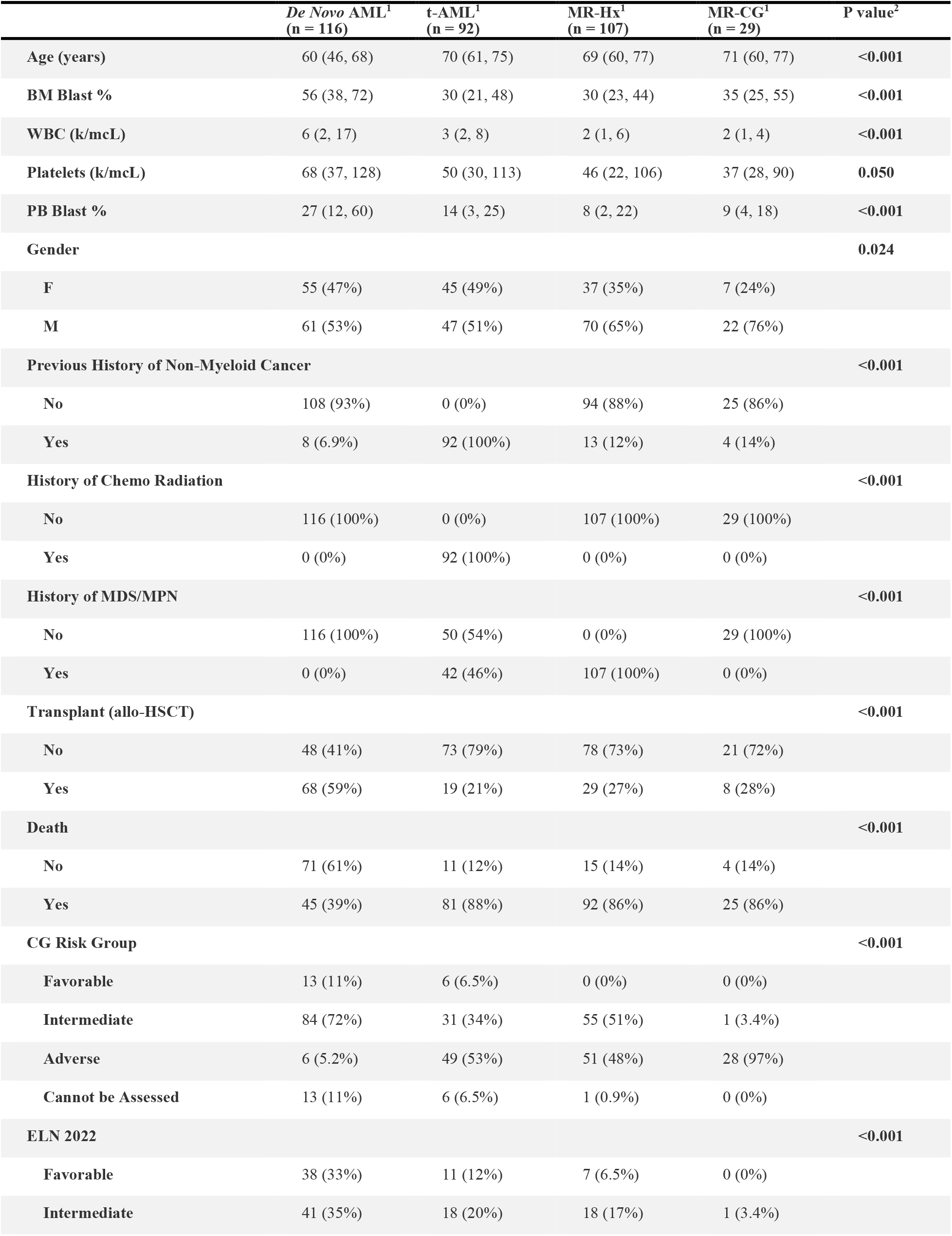
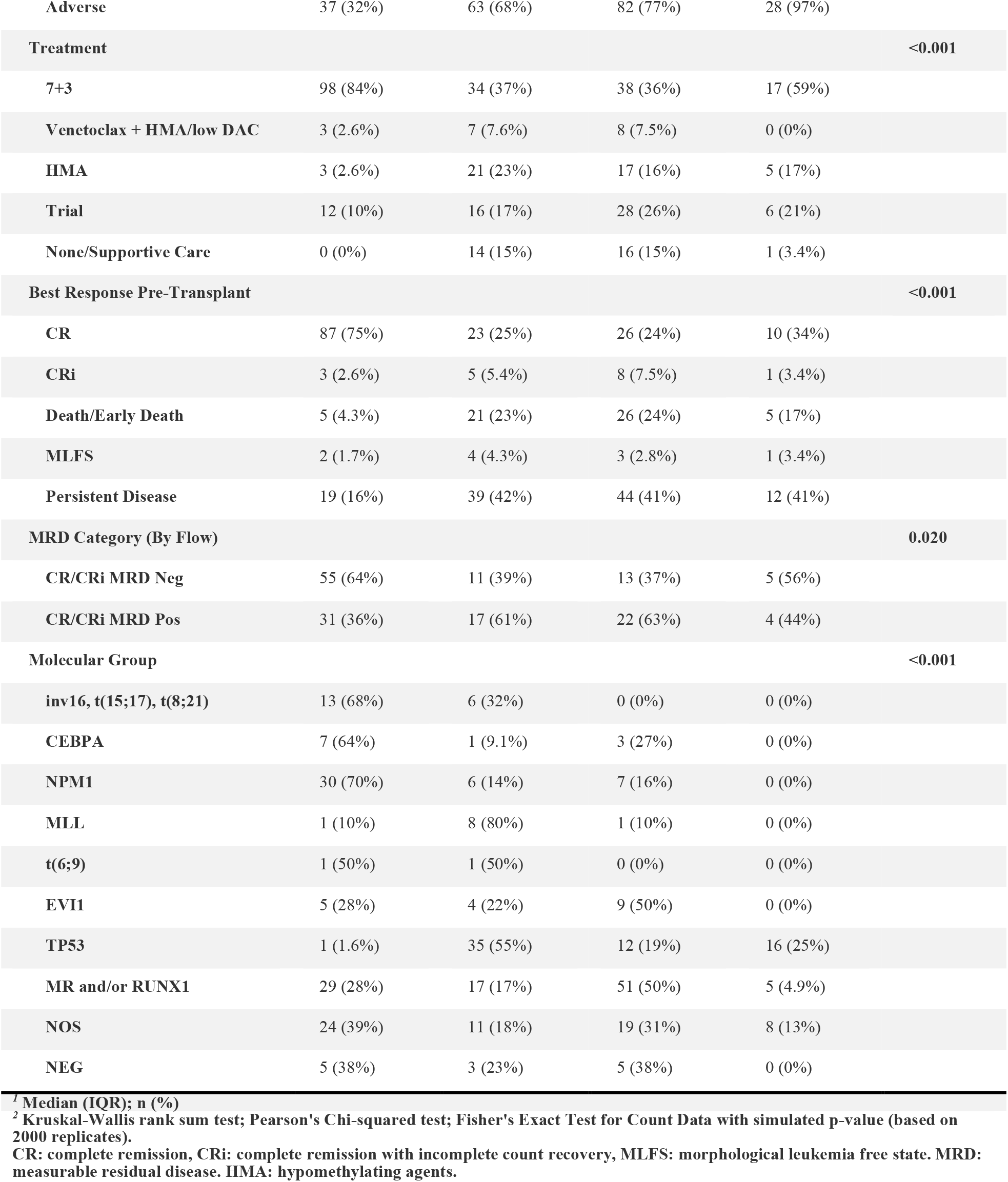
Clinical characteristics of AML ontogeny subtypes.

Gene mutations covered by all 3 panels and additional MR genes (*BCOR, SRSF2, STAG2, U2AF1, ZRSR2*) only covered by RDTB and IMPACT-heme panels were shown in **Figure 1A-B**. As expected, mutations in *NPM1, DNMT3A, IDH1, IDH2* and *FLT3* were predictive for *de novo* AML, while *TP53* mutations were highly predictive for t-AML (**Figure 1B** and **supplemental Figure 1A**). We also confirmed that mutations in *ASXL1, SF3B1, SRSF2, EZH2* and *RUNX1* were predictive for MR-Hx over *de novo* AML^17,28^(**Figure 1B-C** and **supplemental Figure 1B**). Surprisingly, *KRAS* mutations were mostly present in MR-Hx. Both WHO-MR and ICC-MR gene combinations were also predictive for MR-Hx (**Figure 1C**). When t-AML and MR-Hx were compared, *TP53* mutations were more predictive for t-AML while mutations in *SF3B1, SRSF2*, and *RUNX1* were predictive for MR-Hx (**supplemental Figure 1C**). Of note, the mutational profile of MR-CG resembled that of t-AML with a high prevalence of *TP53* mutations (16/29, 55.2% in MR-CG and 36/92, 39.1% in t-AML) and complex karyotype (CK), suggestive of shared biology driven by *TP53* mutations. *TP53* mutations were only present in a small subset of MR-Hx (13/107, 12.1%), and rarely in *de novo* AML (2/116, 1.7%).

**Figure 1.**
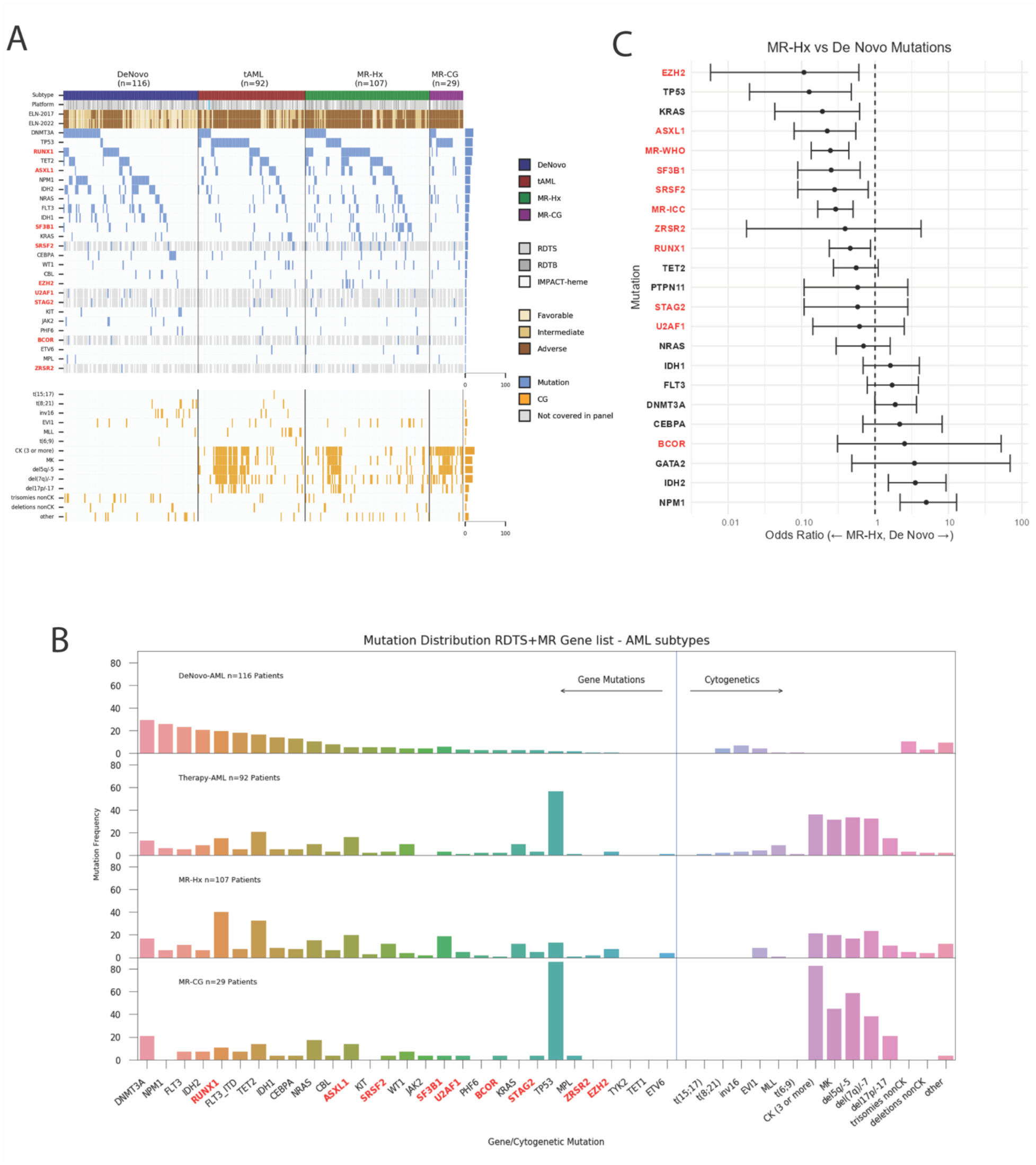
Genomic profile predicts AML ontogeny. A. Oncoplot of AML subtypes (*de novo* AML, t-AML, MR-Hx and MR-CG). NGS panels are indicated as platform (RDTS, RDTB and IMPACT-heme). Genes not covered by RDTS are indicated by grey color. ELN2017 and ELN2022 risk groups are listed. EVI1 indicates EVI1 rearrangements. MLL indicates MLL rearrangements. CK: complex karyotype. MK: monosomal karyotype. B. Bar plots of genomic aberrations in each AML subtypes. Proportions are shown. MR/RUNX1 genes are bolded. C. Association between individual gene mutations and AML ontogeny. Odds ratio was depicted on a log_10_ scale. The comparison is between MR-Hx and de novo AML.

### Correlation between AML ontogeny and genomic classification

Based on previous studies and the new WHO and IC classification^15,18,19^, AML patients were divided into 8 genomic classes: favorable fusions [t(15;17), t(8;21) or inv(16), n=19, 5.5%], *MLL* rearranged (n=10, 2.9%), t(6;9) (n=2, 0.6%), *EVI1* rearranged (n=18, 5.2%), *NPM1* mutations (n=43, 12.5%), *CEBPA bZIP* mutations (n=11, 3.2%), *TP53* mutations (n=64, 18.6%), and *MR/RUNX1* mutations (n=102, 29.7%) (**Table 2** and **supplemental Table 4**). Sixty-two (18%) patients had mutations that did not belong to any of these subgroups (not otherwise specified, NOS). Thirteen patients (3.8%) had no mutations detected (negative, NEG).

Patients with favorable fusions and *CEBPA bZIP* mutations were younger than other subgroups (**supplemental Table 4**). *NPM1* mutated and *MLL* rearranged AML showed female preponderance. The majority of AML with favorable fusions, *CEBPA bZIP* or *NPM1* mutations were *de novo* and a small proportion t-AML (**Figure 2**). Interestingly, 3/11 (27.3%) *CEBPA bZIP* and 7/43 (16.3%) *NPM1* mutated AML had a history of MDS (one had MDS/MPN) with a median interval of 6 months (IQR 3.8-10.5 months) before progressing to AML (**supplemental Table 5**). Among them, 2 *CEBPA bZIP* and 2 *NPM1* mutated patients had MDS-EB2 (10-15% blasts). In addition, 1/10 (10%) *MLL* and 9/18 (50%) *EVI1* rearranged AML had antecedent MDS or MDS/MPN with a median interval of 12 months (IQR 4-24 months) prior to AML **(supplemental Table 5)**. 6 patients were evaluated for *EVI1* rearrangements at the stage of MDS and 5 were positive, 4 with 10-15% blasts and 1 with 5%. Of note, nearly all these patients would be classified as *de novo* AML based on the WHO^2022^ and approximately 50% would if ICC^2022^ is applied.

**Figure 2.**
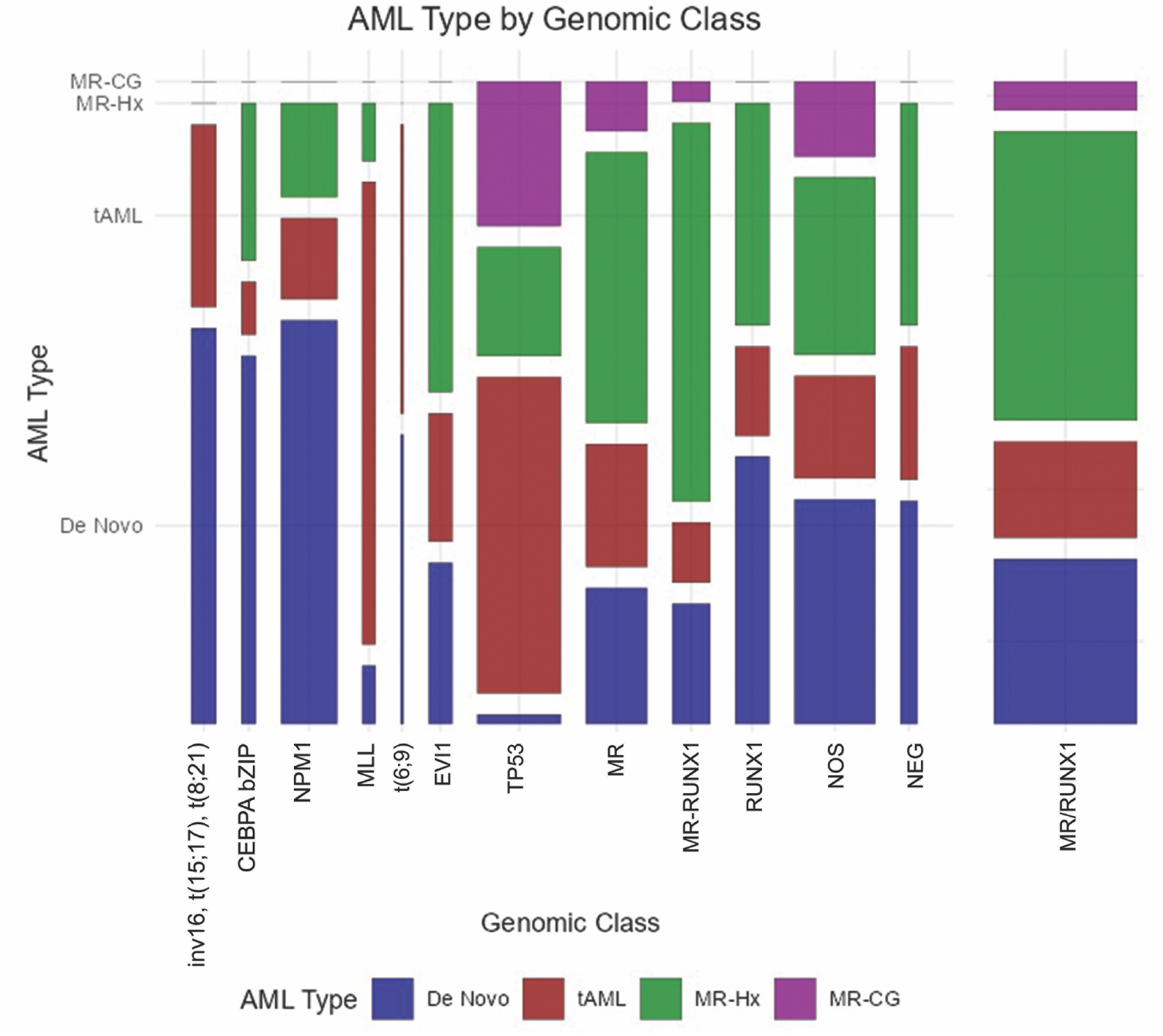
Distribution of AML ontogeny subtypes in each genomic class. The width of each bar represents the number of patients. MR indicates AML patients with MR gene mutations (including *ASXL1, BCOR, EZH2, STAG2, SF3B1, SRSF2, ZRSR2, U2AF1*, but no *RUNX1* mutations). *MR-RUNX1* indicates AML patients with both MR gene and *RUNX1* gene mutations. *RUNX1* indicates AML patients with *RUNX1* but no MR gene mutations. MR or *RUNX1* indicates AML patients with MR and/or *RUNX1* gene mutations. NOS indicates AML patients with mutations detected but unable to assign to a well-defined entity. NEG indicates AML patients with no mutations or rearrangements detected.

*TP53* mutated AML comprised predominantly t-AML (35/64, 54.7%), MR-CG (16/64, 25%) and MR-Hx (12/64, 18.8%) groups. Only 1/64 (1.6%) *TP53* mutated AML was *de novo* (**supplemental Table 4**). This patient had *de novo* monocytic AML with *TET2* and TP53 mutations and extensive necrosis in the marrow. FISH studies performed on peripheral blood samples were negative. The patient had refractory disease and died in 6 months. 31/64 (48.4%) patients with *TP53* mutated AML had a prior MDS (19 t-MDS) or MDS/MPN stage and the median interval was 11 months (IQR 4-15 months) before the development of AML. Strikingly, 61/64 (95.3%) patients with *TP53* mutated AML had CG abnormalities involving -5/del5q, -7/del7q, -17/del17p and/or CK. Conversely, 61/119 (51.3%) patients with such CG abnormalities harbored *TP53* mutations. Specifically, 46/55 (83.6%) patients with concurrent CK and monosomal karyotypes harbored *TP53* mutations.

*MR/RUNX1* mutated AML were mostly MR-Hx (51/102, 50%), and t-AML (18/102, 17%) (**Figure 2** and **supplemental Table 4**). Only 5 patients were MR-CG (4.9%). Importantly, 29/102 (28%) patients were *de novo* AML. Of note, 10 (9.6%) patients had isolated trisomy 8 and/or del20q only, which was newly added by ICC but not WHO as MDS-defining CG abnormalities, and 5 of them were *de novo* AML (**supplemental Table 6**).

### AML ontogeny predicts outcome independent of ELN2022 risk stratification

*De novo* AML had a significantly better overall survival (OS) than all 3 other subtypes (median: not reached vs. 8.4 months (95% CI: 6-12.1 months) in t-AML, 9.4 months (95% CI: 6.5-14.7 months) in MR-Hx and 7.7 months (95% CI: 4.3-17.1 months) in MR-CG, p<0.0001, **Figure 3A**). This difference remained statistically significant in patients treated with daunorubicin+cytarabine (7+3) induction therapy, if *TP53* mutated patients were excluded or after stratified by ELN2022 risk groups, genomic classes, CG risks and NGS platforms (**supplemental Figures 2-3)**. AML ontogeny further stratifies the outcomes of all 3 ELN2022 risk groups but of only the intermediate (not favorable, or adverse) CG risk group (**supplemental Figure 4**).

**Figure 3.**
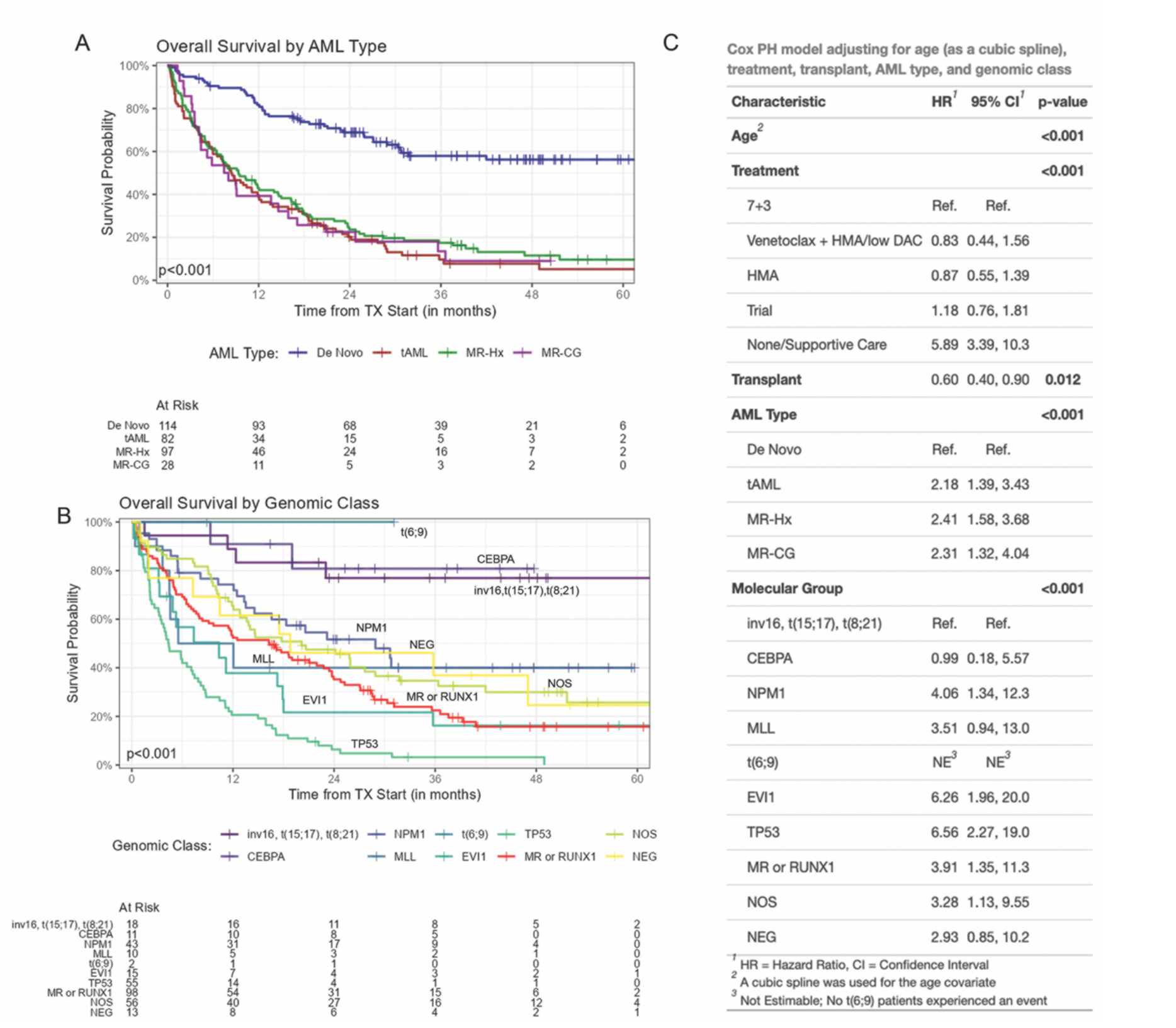
Both AML ontogeny and ELN risks have independent prognostic values. A. Kaplan-Meier curves of overall survival divided by AML ontogeny subtypes. B. Kaplan-Meier curves of overall survival divided by genomic classes. Both patients with t(6;9) received allo-HSCT. C. AML ontogeny related risk was evaluated in multivariable Cox-regression models adjusting for age (modeled by cubic spline), initial treatment at diagnosis, allogenic transplant (modeled as time-dependent variable), and genomic classes. 7+3, daunorubicin+cytarabine including CPX-351; HMA, hypomethylating agents; low DAC, low-dose cytarabine.

Genomic classes significantly correlated with OS (**Fig 3B**). The impact of genomic class was more significant in *de novo* AML and only borderline significant in t-AML (**supplemental Figure 5**). Patients with favorable fusions and *CEBPA* mutations had the most favorable OS (median not reached, **Fig 3B**). *NPM1* mutated and *MR/RUNX1* mutated AML had a median OS of 28.9 months (95% CI: 14.6 months-not reached) and 16.3 months (95% CI: 9.1-23.4 months), respectively. Interestingly, the outcome of both *CEBPA* and *NPM1* mutated AML were further stratified by ontogeny (**supplemental Figure 6B-C**), somewhat in contrast to a recent large cohort showing *NPM1* mutated t-AML has similar OS to *de novo* counterpart after excluding early death ^29^. As expected, either *TP53* mutated or *EVI1* rearranged AML had dismal outcome (median OS: 4.4 months (95% CI: 3.4-7.5 months) and 10.3 months (95% CI: 4.9-35.7 months), respectively) regardless of ontogeny (**supplemental Figure 6D-E**). Further analysis on *TP53* mutated AML showed that neither mutation burden, biallelic status, complex karyotypes, or co-mutations in signaling molecules affected the outcomes (**supplemental Figure 7**).

In univariate analysis, *CEBPA, DNMT3A, NPM1* and *IDH2* mutations were associated with a superior OS (**supplemental Figure 8A**). In contrast, *TP53, KRAS, NOTCH2, EZH2* and *SF3B1* mutations were associated with an inferior OS. Of note, mutations in most MR genes and *RUNX1* were not significantly associated with outcome. To study the interaction between mutations and ontogeny, the impact of mutations on OS was adjusted for AML subtype. To this end, the prognostic values of many gene mutations (*DNMT3A, NPM1, IDH2, KRAS, NOTCH2, EZH2* and *SF3B1*) were no longer significant, while *TP53* and *CEBPA* mutations remained prognostic (**supplemental Figure 8B**). In a multivariate analysis adjusting for age, initial treatment for induction, and allo-HSCT, both AML subtypes and genomic classes or ELN2022 risks were significantly associated with outcomes (**Figure 3C and supplemental Figure 8C**), demonstrating that ontogeny and genomics provide independent prognostic values.

### Specificity of myelodysplasia related gene mutations for MR-Hx AML

We next focused on *MR/RUNX1* mutated AML. Although a previous study had suggested that MR gene mutations are highly specific to MR-Hx (with 95% specificity)^17^, the specificity of these gene mutations to MR-Hx remains controversial ^28,30^. We decided to address these issues with our well-annotated cohort. We first examined the *MR/RUNX1* group that were sequenced by targeted NGS panels covering all 8 MR and *RUNX1* genes. Among 42 patients, mutations of *MR/RUNX1* genes were present in 14 *de novo* AML and 28 MR-Hx AML, resulting in a positive predictive value of only 66% for MR-Hx. This is similar to our entire cohort (positive predictive value 51/80, 64%, specificity 74%) regardless of NGS panels used (**Figure 2**) as well as other studies^28,30^. If *RUNX1* mutations are excluded, the positive predictive value and specificity of MR gene mutations for MR-Hx is 70% and 75%, respectively.

### *De novo* AML with myelodysplasia related gene mutations do not have an adverse outcome

Nearly one third of the AML patients with *MR/RUNX1* mutations were *de novo*. The ELN2022 guideline risk stratifies these patients to the adverse-risk group regardless of ontogeny. We thus investigated the impact of ontogeny on the OS in the patients carrying these mutations. In this group, there were very few patients with MR-CG, insufficient for a statistical analysis. However, patients with *de novo* AML had a remarkably better outcome than those with MR-Hx or t-AML (median OS: not reached vs 7.3 (95% CI: 5.1-17 months) or 6 months (95% CI: 3 months-not reached), p<0.001, **Figure 4A, and supplemental Table 7**). This was also true in the patients with MR only or *RUNX1* only mutations (**supplemental Figure 9 A-B**). Comparable OS was observed between ELN2017 intermediate and adverse risk groups, suggesting even *ASXL1* and *RUNX1* mutations may not be able to risk stratify this patient population (**supplemental Figure 9C**). The proportion of adverse CG abnormalities was significantly lower in *de novo* AML subgroups (**supplemental Table 7**), but the difference in OS was small between intermediate and adverse CG risk groups (**supplemental Figure 9D**). Univariate analysis stratified by AML subtypes showed mutations in *NRAS* and *KRAS* but not any of the individual *MR/RUNX1* genes were associated with an inferior OS (**supplemental Figure 10A**). A multivariate analysis showed that age, initial treatment for induction, allo-HSCT, and AML ontogeny but not *NRAS* or *KRAS* mutations or CG risks remained statistically significant **(supplemental Figure 10B)**. The prognostic values of AML ontogeny were even more significant if the analysis was restricted to the patients uniformly treated with induction chemotherapy with daunorubicin+cytarabine regimen (**Figure 4B**). *De novo* AML with *MR/RUNX1* mutations had an outcome that fell in between favorable- and intermediate-risk groups when compared to the remainder of the patients stratified by either ELN 2017 or ELN2022 risks (**Figure 4C and supplemental Figure 11**). Interestingly, *de novo* AML with WHO-MR mutations had an outcome overlapping with ELN2022 intermediate-risk group but *de novo* AML with only *RUNX1* mutations had a rather favorable outcome (**supplemental Figure 11D**).

**Figure 4.**
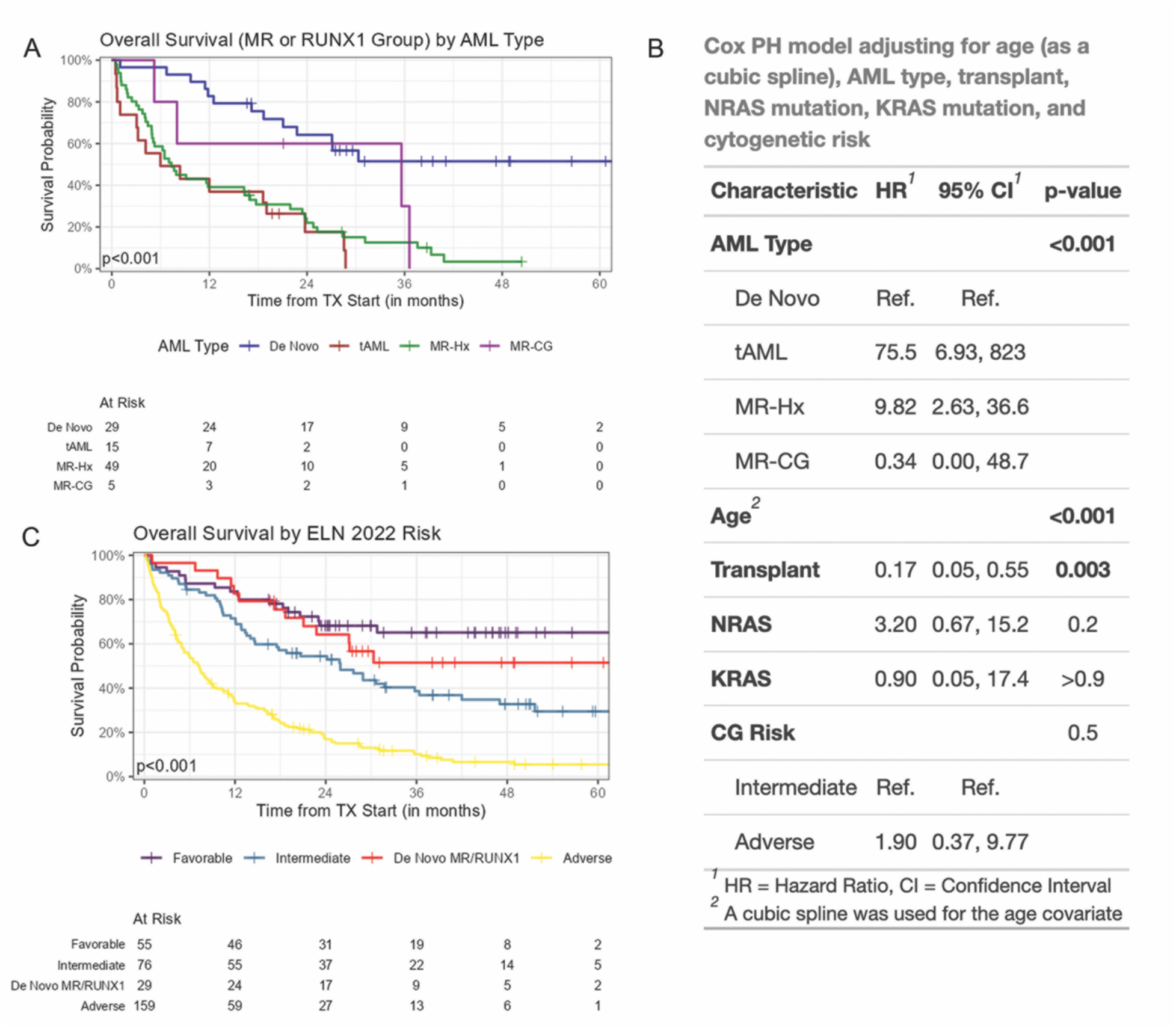
AML ontogeny determines the outcome of AML with *MR/RUNX1* gene mutations. A. Kaplan-Meier curves of overall survival of AML with *MR/RUNX1* gene mutations divided by ontogeny. B. AML ontogeny related risk was evaluated in multivariable Cox-regression models adjusting for age (modeled by cubic spline), allogenic transplant (modeled as time-dependent variable), *NRAS/KRAS* mutations, and CG risks in AML patients with *MR/RUNX1* mutations uniformly treated with 7+3 induction therapy. C. Kaplan-Meier curves of AML patients were divided by ELN2022 risk groups. *De novo* AML with *MR/RUNX1* gene mutations separated from ELN2022 adverse group show an outcome falling in between favorable and intermediate risk groups.

## Discussion

The most striking finding of our study is the inaccuracy of ontogeny assignment based on initial database registry, which is likely not unique to this study and/or our cancer center. It may also have been encountered by many large cohorts of AML studies as several landmark genomic studies on *de novo* AML included at least 15-20% cases harboring apparent MDS-defining cytogenetic abnormalities with or without *TP53* mutations ^13,17,31-35^, consistent with either t-AML or AML-MRC^WHO2016^. This number is certainly an underestimate as not all t-AML or AML-MRC^WHO2016^ cases have MDS-defining cytogenetic abnormalities. The inaccurate assignment of ontogeny is due to 1) inadequate history documentation of antecedent MDS or MDS/MPN, or cytotoxic treatment for other types of malignancies/disorders; 2) delayed and/or overlooked cytogenetic information showing MDS-defining abnormalities; and 3) overcalling AML-MRC^WHO2016^ solely based on morphologic dysplasia. Our study emphasizes the importance of cohort annotation to render an accurate assignment of AML ontogeny.

Our study demonstrates the prognostic value of AML ontogeny independent of genomics and/or ELN risk stratification. The designation of ontogeny as either MR-Hx/MR-CG or t-AML has clinical implications. Many studies have shown that “secondary” AML, mostly MR-Hx/MR-CG and t-AML, is associated with an inferior outcome with or without allo-HSCT ^16,33,34,36-40^. Our study has confirmed these findings by showing MR-Hx, MR-CG and t-AML all confer worse outcomes than de novo AML even after adjusting for age, treatment, transplant, and ELN risks. Importantly, several clinical trials had shown that CPX-351, a liposomal form of daunorubicin+cytarabine, improves outcomes of AML-MRC^WHO2016^ and t-AML compared to conventional daunorubicin+cytarabine^41-43^. Therefore, an accurate designation of AML ontogeny remains important for clinical decisions.

How to best incorporate ontogeny and genomics into AML classification and risk stratification to guide clinical management remains to be determined particularly for AML with MR gene mutations. Although many MR genes are moderately predictive for MR-Hx, our study found that they are also frequent in *de novo* AML resulting in only an approximate 70% specificity to MR-Hx, significantly lower than the 95% previously reported ^17^. The discrepancy may be due to different cohorts but is more likely related to misclassification. The suboptimal specificity of MR gene mutations has also been observed in a few recent studies ^28,30^. This questions the approach of classifying these MR genes into one entity largely aiming to replace MR-Hx or AML-MRC^WHO2016^.

Several studies have demonstrated inferior outcomes in AML with MR gene mutations, but its independent prognostic value in *de novo* AML has been controversial ^21-23,44^. Recent studies have shown that MR gene mutations in a favorable-risk group (mostly *de novo*) do not affect outcome^30,45^. Our findings indicate that the outcome of AML with MR gene mutations in ELN2017 intermediate/adverse risk group is stratified by ontogeny and *de novo* patients had an OS not inferior to intermediate risk group. Therefore, it would be reasonable to restrict the diagnostic criteria of AML-MR to: 1) history of MDS or MDS/MPN (i.e. MR-Hx); or 2) MDS-defining CGA (i.e. MR-CG). A majority of patients with AML with MR gene mutations will meet one or both of these two criteria and therefore be classified as AML-MR. In order to better study the biology and management of *de novo* AML with MR gene mutations that do not meet either one of these two criteria, a provisional entity could be established: (*de novo*) AML with MR gene mutations. Our data demonstrate that assignment of mutations in all these MR genes into adverse risk groups is likely oversimplified and inaccurate and may lead to mismanagement in a subset of these patients. Our data also suggest that ontogeny of MR-Hx and t-AML bears more prognostic weight than MR gene mutations and should be included in AML risk stratification for the patients lacking favorable risk factors. The findings that ontogeny carries prognostic value independent of genomics are intriguing, which may be attributed to clonal architectural complexity, immune dysregulations, and microenvironment among other factors.

Our findings support the classification of AML with *TP53* mutations as a distinct entity due to its dismal prognosis regardless of ontogeny, which has recently been demonstrated by several studies ^35,37,46,47^. We and others show that nearly >90% of AML with *TP53* mutations harbor CK or monosomal karyotype involving chromosomes 5, 7 and/or 17^46^. Notably, the similar mutational profile between MR-CG and t-AML is largely driven by *TP53* mutations ^47-49^, further supporting a separation of AML with *TP53* mutations from others.

Because this is a retrospective, single-centered study at a tertiary cancer center, there is a potential inherent bias towards more adverse risk patients. However, we were able to review the detailed clinical history and pathologic findings of every patient in this cohort. To our knowledge, this is to date the most well-annotated cohort with a large number of patients with therapy related or antecedent MDS history, which is essential for addressing the clinical impact of ontogeny. Although not all the patients were uniformly treated, most of the patients received standard intensive induction chemotherapy with the daunorubicin+cytarabine regimen and the conclusion remained valid when analysis was restricted to this subset of patients. Moreover, preliminary data from several recent studies performed in different centers support our findings with respect to AML with MR gene mutations^50-53^, arguing against the inclusion of all MR gene mutations into the adverse-risk group. A recent study suggested that the number of MR mutations predicts outcome^16^. Unfortunately, our study was not powered to validate the results due to an incomplete coverage of all MR genes in a subset of patients. More studies are urgently needed to delineate the role of individual MR genes, its combination and allelic burden in AML classification and risk stratification^54,55^.

## Data Availability

All data produced in the present work are contained in the manuscript

## Abbreviations

WHO: World Health Organization
ICC: International Consensus Classification
ELN: European LeukemiaNet
AML-MRC: Acute myeloid leukemia with myelodysplasia-related changes
t-AML: therapy-related acute myeloid leukemia
MDS: myelodysplastic syndromes
MDS/MPN: myelodysplastic/myeloproliferative syndromes
MR-Hx: acute myeloid leukemia, myelodysplasia-related based on history of MDS or MDS/MPN
MR-CG: acute myeloid leukemia, myelodysplasia-related based on MDS-defining cytogenetic changes (no history of MDS or MDS/MPN)

## Acknowledgement

We thank Michael R. Waarts for his critical reading. This study was funded by the Center for Hematologic Malignancies at MSKCC and in part through the NIH/NCI Cancer Center Support Grant P30 CA008748. A.J.S. is supported by a Young Investigator Award from the Edward P. Evans Foundation. A.J.D. is a William Raveis Charitable Fund Physician-Scientist of the Damon Runyon Cancer Research Foundation (PST-24-19). S.F.C. is supported by a National Cancer Institute grant (K08 CA241371-01A1). J.L.G. is supported by a National Cancer Institute grant (NCI K08CA230172) and Equinox Cycle for Survival. O.A.-W. is supported in part by the Edward P. Evans Foundation, NIH/NCI (R01 CA251138 and R01 CA242020), NIH/NHLBI (R01 HL128239), NIH/NCI P50 CA254838-01, and the Leukemia & Lymphoma Society. R.L.L. is supported by a Cycle For Survival Innovation Grant and National Cancer Institute R35 CA197594. WX is supported by Alex’s Lemonade Stand Foundation and the Runx1 Research Program, MSK’s Cycle for Survival’s Equinox Innovation Award in Rare Cancers, MSK Leukemia SPORE Career Enhancement Program and a National Cancer Institute grant (K08CA267058-01).

## Authorship contributions

J.G.M. performed computational analysis and curated mutations. D.N. and A. K. performed statistical analysis. C.F., Z.S.S., J.C., B.B., Z.D.E., A.D.G., M.R. and W.X. performed chart review. N.F. A.S.M. and M.E.A. assisted mutation analysis. A.J.S, A.D., S.F.C., J.L.G, M.B.G., R.K.R., E.B, O.I.A., E.M.S., A.D.G., M.S.T. and R.L.L provided patient care. E.P. supervised computational analysis. Y.Z. reviewed cytogenetic data. M.R. and W.X. reviewed pathology. W.X. initiated, designed and supervised the study. W.X. drafted and all authors finalized and approved the manuscript.

## Conflict of Interest

M.E.A served as consultant for Janssen Global Services, Bristol-Myers Squibb, AstraZeneca, and Roche; and has received honoraria from Biocartis, Invivoscribe, physician educational resources (PER), Peerview Institute for medical education, clinical care options, RMEI medical education. A.J.S. reports his spouse is an employee of Bristol-Myers Squibb. B.J.B served on the advisory board for Oncovalent and Bristol Myers Squibb. S.F.C. is a consultant for and holds equity interest in Imago Biosciences, none of which are directly related to the content of this paper. J.L.G. received consulting fees from GLG. M.B.G. receives research support from Actinium, Amgen, and Sanofi and has served in an advisory role for Sanofi, Novartis, and Allogene. R.K.R has received consulting Fees from Incyte Corporation, Celgene/BMS, Blueprint, Abbvie, CTI, Stemline, Galecto, Pharmaessentia, Constellation/Morphosys, Sierra Oncology/GSK, Sumitomo Dainippon Kartos, Servier, Zentalis, Karyopharm and research funding from Constellation pharmaceuticals, Ryvu, Zentalis and Stemline Therapeutics. O.A.-W. has served as a consultant for H3B Biomedicine, Foundation Medicine Inc., Merck, Prelude Therapeutics, and Janssen, and is on the Scientific Advisory Board of Envisagenics Inc., AIChemy, Harmonic Discovery Inc., and Pfizer Boulder; O.A.-W. has received prior research funding from H3B Biomedicine, Nurix Therapeutics, Minovia Therapeutics, and LOXO Oncology unrelated to the current manuscript. A.D.G. received research funding from Celularity, ADC Therapeutics, Aprea, AROG, Pfizer, Prelude, and Trillium; received research funding from and served as a consultant for Aptose and Daiichi Sankyo; served as a consultant and member of advisory committees for Astellas, Celgene, and Genentech; received research funding from, served as a consultant for, and was a member of advisory committees for AbbVie; and received honoraria from Dava Oncology. M.S.T. has received research funding from AbbVie, Orsenix, BioSight, Glycomimetics, Rafael Pharmaceuticals and Amgen. M.S.T. is on the advisory boards of AbbVie, Daiichi-Sankyo, Orsenix, KAHR, Jazz Pharmaceuticals, Roche, BioSight, Novartis, Innate Pharmaceuticals, Kura, Syros Pharmaceuticals and Ipsen Biopharmaceuticals. M.S.T. has received royalties from UpToDate. M.S.T. is on DSMB of HOVON protocol Ho156 and adjudication committee of Foghorn protocol FHD-286. RLL is on the supervisory board of Qiagen and is a scientific advisor to Imago, Mission Bio, Syndax. Zentalis, Ajax, Bakx, Auron, Prelude, C4 Therapeutics and Isoplexis for which he receives equity support. RLL receives research support from Ajax and Abbvie and has consulted for Incyte, Janssen, Morphosys and Novartis. He has received honoraria from Astra Zeneca and Kura for invited lectures and from Gilead for grant reviews. E.P is a founder, equity holder and holds fiduciary role in Isabl Inc. M.R. is on the scientific advisory board in Auron Pharmaceutical for which he received equity support. He receives research funding from Celularity, Roche-Genentech, Beat AML and NGM and travel fund from BD Biosciences. W.X. has received research support from Stemline Therapeutics.

## Notes

### Author Declarations

the Institutional Review Board at the memorial sloan kettering cancer center had approved this study.

## Reference

1. Pollyea DA, Bixby D, Perl A, et al. NCCN Guidelines Insights: Acute Myeloid Leukemia, Version 2.2021. J Natl Compr Canc Netw. 2021;19(1):16–27.

2. Döhner H, Wei AH, Appelbaum FR, et al. Diagnosis and management of AML in adults: 2022 recommendations from an international expert panel on behalf of the ELN. Blood. 2022;140(12):1345–1377.

3. Bennett JM, Catovsky D, Daniel M-T, et al. Proposals for the Classification of the Acute Leukaemias French-American-British (FAB) Co-operative Group. British Journal of Haematology. 1976;33(4):451–458.

4. Arber DA, Orazi A, Hasserjian R, et al. The 2016 revision to the World Health Organization classification of myeloid neoplasms and acute leukemia. Blood. 2016;127(20):2391–2405.

5. Vardiman JW, Thiele J, Arber DA, et al. The 2008 revision of the World Health Organization (WHO) classification of myeloid neoplasms and acute leukemia: rationale and important changes. Blood. 2009;114(5):937–951.

6. Döhner H, Estey E, Grimwade D, et al. Diagnosis and management of AML in adults: 2017 ELN recommendations from an international expert panel. Blood. 2017;129(4):424–447.

7. Weinberg OK, Pozdnyakova O, Campigotto F, et al. Reproducibility and prognostic significance of morphologic dysplasia in de novo acute myeloid leukemia. Modern Pathology. 2015;28(7):965–976.

8. Weinberg OK, Gibson CJ, Blonquist TM, et al. NPM1 mutation but not RUNX1 mutation or multilineage dysplasia defines a prognostic subgroup within de novo acute myeloid leukemia lacking recurrent cytogenetic abnormalities in the revised 2016 WHO classification. Am J Hematol. 2017;92(7):E123–e124.

9. Wandt H, Schäkel U, Kroschinsky F, et al. MLD according to the WHO classification in AML has no correlation with age and no independent prognostic relevance as analyzed in 1766 patients. Blood. 2008;111(4):1855–1861.

10. Miesner M, Haferlach C, Bacher U, et al. Multilineage dysplasia (MLD) in acute myeloid leukemia (AML) correlates with MDS-related cytogenetic abnormalities and a prior history of MDS or MDS/MPN but has no independent prognostic relevance: a comparison of 408 cases classified as “AML not otherwise specified” (AML-NOS) or “AML with myelodysplasia-related changes” (AML-MRC). Blood. 2010;116(15):2742–2751.

11. Haferlach T, Schoch C, Löffler H, et al. Morphologic dysplasia in de novo acute myeloid leukemia (AML) is related to unfavorable cytogenetics but has no independent prognostic relevance under the conditions of intensive induction therapy: results of a multiparameter analysis from the German AML Cooperative Group studies. J Clin Oncol. 2003;21(2):256–265.

12. Tyner JW, Tognon CE, Bottomly D, et al. Functional genomic landscape of acute myeloid leukaemia. Nature. 2018;562(7728):526–531.

13. Genomic and Epigenomic Landscapes of Adult De Novo Acute Myeloid Leukemia. New England Journal of Medicine. 2013;368(22):2059–2074.

14. Patel JP, Gönen M, Figueroa ME, et al. Prognostic Relevance of Integrated Genetic Profiling in Acute Myeloid Leukemia. New England Journal of Medicine. 2012;366(12):1079–1089.

15. Papaemmanuil E, Gerstung M, Bullinger L, et al. Genomic Classification and Prognosis in Acute Myeloid Leukemia. N Engl J Med. 2016;374(23):2209–2221.

16. Tazi Y, Arango-Ossa JE, Zhou Y, et al. Unified classification and risk-stratification in Acute Myeloid Leukemia. Nat Commun. 2022;13(1):4622.

17. Lindsley RC, Mar BG, Mazzola E, et al. Acute myeloid leukemia ontogeny is defined by distinct somatic mutations. Blood. 2015;125(9):1367–1376.

18. Khoury JD, Solary E, Abla O, et al. The 5th edition of the World Health Organization Classification of Haematolymphoid Tumours: Myeloid and Histiocytic/Dendritic Neoplasms. Leukemia. 2022;36(7):1703–1719.

19. Arber DA, Orazi A, Hasserjian RP, et al. International Consensus Classification of Myeloid Neoplasms and Acute Leukemias: integrating morphologic, clinical, and genomic data. Blood. 2022;140(11):1200–1228.

20. Ohgami RS, Ma L, Merker JD, et al. Next-generation sequencing of acute myeloid leukemia identifies the significance of TP53, U2AF1, ASXL1, and TET2 mutations. Mod Pathol. 2015;28(5):706–714.

21. Song GY, Kim T, Ahn SY, et al. Allogeneic hematopoietic cell transplantation can overcome the adverse prognosis indicated by secondary-type mutations in de novo acute myeloid leukemia. Bone Marrow Transplant. 2022;57(12):1810–1819.

22. Caprioli C, Lussana F, Salmoiraghi S, et al. Clinical significance of chromatin-spliceosome acute myeloid leukemia: a report from the Northern Italy Leukemia Group (NILG) randomized trial 02/06. Haematologica. 2021;106(10):2578–2587.

23. Gardin C, Pautas C, Fournier E, et al. Added prognostic value of secondary AML-like gene mutations in ELN intermediate-risk older AML: ALFA-1200 study results. Blood Adv. 2020;4(9):1942–1949.

24. Xiao W, Bharadwaj M, Levine M, et al. PHF6 and DNMT3A mutations are enriched in distinct subgroups of mixed phenotype acute leukemia with T-lineage differentiation. Blood Adv. 2018;2(23):3526–3539.

25. Forbes SA, Beare D, Boutselakis H, et al. COSMIC: somatic cancer genetics at high-resolution. Nucleic Acids Res. 2017;45(D1):D777–d783.

26. Chakravarty D, Gao J, Phillips SM, et al. OncoKB: A Precision Oncology Knowledge Base. JCO Precis Oncol. 2017;2017.

27. Karczewski KJ, Francioli LC, Tiao G, et al. The mutational constraint spectrum quantified from variation in 141,456 humans. Nature. 2020;581(7809):434–443.

28. Gao Y, Jia M, Mao Y, et al. Distinct Mutation Landscapes Between Acute Myeloid Leukemia With Myelodysplasia-Related Changes and De Novo Acute Myeloid Leukemia. Am J Clin Pathol. 2022;157(5):691–700.

29. Othman J, Meggendorfer M, Tiacci E, et al. Overlapping features of therapy-related and de novoNPM1-mutated AML. Blood. 2022.

30. Lachowiez CA, Long N, Saultz JN, et al. Comparison and validation of the 2022 European LeukemiaNet guidelines in acute myeloid leukemia. Blood Adv. 2022.

31. Schoch C, Kern W, Schnittger S, Hiddemann W, Haferlach T. Karyotype is an independent prognostic parameter in therapy-related acute myeloid leukemia (t-AML): an analysis of 93 patients with t-AML in comparison to 1091 patients with de novo AML. Leukemia. 2004;18(1):120–125.

32. van der Werf I, Wojtuszkiewicz A, Meggendorfer M, et al. Splicing factor gene mutations in acute myeloid leukemia offer additive value if incorporated in current risk classification. Blood Adv. 2021;5(17):3254–3265.

33. Martínez-Cuadrón D, Megías-Vericat JE, Serrano J, et al. Treatment patterns and outcomes of 2310 patients with secondary acute myeloid leukemia: a PETHEMA registry study. Blood Adv. 2022;6(4):1278–1295.

34. Lalayanni C, Gavriilaki E, Athanasiadou A, et al. Secondary Acute Myeloid Leukemia (sAML): Similarly Dismal Outcomes of AML After an Antecedent Hematologic Disorder and Therapy Related AML. Clin Lymphoma Myeloma Leuk. 2022;22(4):e233–e240.

35. Awada H, Durmaz A, Gurnari C, et al. Machine learning integrates genomic signatures for subclassification beyond primary and secondary acute myeloid leukemia. Blood. 2021;138(19):1885–1895.

36. Kuykendall A, Duployez N, Boissel N, Lancet JE, Welch JS. Acute Myeloid Leukemia: The Good, the Bad, and the Ugly. Am Soc Clin Oncol Educ Book. 2018;38:555–573.

37. Tashakori M, Kadia T, Loghavi S, et al. TP53 copy number and protein expression inform mutation status across risk categories in acute myeloid leukemia. Blood. 2022;140(1):58–72.

38. Granfeldt Østgård LS, Medeiros BC, Sengeløv H, et al. Epidemiology and Clinical Significance of Secondary and Therapy-Related Acute Myeloid Leukemia: A National Population-Based Cohort Study. J Clin Oncol. 2015;33(31):3641–3649.

39. Schmaelter AK, Labopin M, Socié G, et al. Inferior outcome of allogeneic stem cell transplantation for secondary acute myeloid leukemia in first complete remission as compared to de novo acute myeloid leukemia. Blood Cancer J. 2020;10(3):26.

40. Gerstung M, Papaemmanuil E, Martincorena I, et al. Precision oncology for acute myeloid leukemia using a knowledge bank approach. Nat Genet. 2017;49(3):332–340.

41. Lancet JE, Uy GL, Newell LF, et al. CPX-351 versus 7+3 cytarabine and daunorubicin chemotherapy in older adults with newly diagnosed high-risk or secondary acute myeloid leukaemia: 5-year results of a randomised, open-label, multicentre, phase 3 trial. Lancet Haematol. 2021;8(7):e481–e491.

42. Lancet JE, Uy GL, Cortes JE, et al. CPX-351 (cytarabine and daunorubicin) Liposome for Injection Versus Conventional Cytarabine Plus Daunorubicin in Older Patients With Newly Diagnosed Secondary Acute Myeloid Leukemia. J Clin Oncol. 2018;36(26):2684–2692.

43. Price K, Cao Z, Lipkin C, Profant D, Robinson S. Comparison of Hospital Length of Stay and Supportive Care Utilization Between Patients Treated with CPX-351 and 7+3 for Therapy-Related Acute Myeloid Leukemia or Acute Myeloid Leukemia with Myelodysplasia-Related Changes. Clinicoecon Outcomes Res. 2022;14:21–34.

44. Lachowiez CA, Loghavi S, Furudate K, et al. Impact of splicing mutations in acute myeloid leukemia treated with hypomethylating agents combined with venetoclax. Blood Adv. 2021;5(8):2173–2183.

45. Wright MF, Pozdnyakova O, Hasserjian RP, et al. Secondary-type mutations do not impact prognosis in acute myelogenous leukemia AML with mutated NPM1. Am J Hematol. 2022;97(12):E462–e465.

46. Grob T, Al Hinai ASA, Sanders MA, et al. Molecular characterization of mutant TP53 acute myeloid leukemia and high-risk myelodysplastic syndrome. Blood. 2022;139(15):2347–2354.

47. Weinberg OK, Siddon A, Madanat YF, et al. TP53 mutation defines a unique subgroup within complex karyotype de novo and therapy-related MDS/AML. Blood Adv. 2022;6(9):2847–2853.

48. Fuhrmann I, Lenk M, Haferlach T, et al. AML, NOS and AML-MRC as defined by multilineage dysplasia share a common mutation pattern which is distinct from AML-MRC as defined by MDS-related cytogenetics. Leukemia. 2022;36(7):1939–1942.

49. Montalban-Bravo G, Kanagal-Shamanna R, Class CA, et al. Outcomes of acute myeloid leukemia with myelodysplasia related changes depend on diagnostic criteria and therapy. Am J Hematol. 2020;95(6):612–622.

50. Jentzsch M, Bischof L, Ussmann J, et al. Prognostic Impact of the 2022 European Leukemia Net Risk Classification in Patients with Acute Myeloid Leukemia Undergoing Allogeneic Stem Cell Transplantation. Blood. 2022;140(Supplement 1):10601–10602.

51. Alkaabba F, Williams M, Corley E, et al. Prognostication of the 2016-Who AML with Myelodysplasia-Related Changes Subclass and Mutations in MDS-Related Genes Using the 2022 European Leukemianet Risk Classification: A Retrospective Study. Blood. 2022;140(Supplement 1):3237–3238.

52. Senapati J, Short N, Maiti A, et al. Splicing Gene Mutations Do Not Affect Outcomes in Newly Diagnosed Patients with Acute Myeloid Leukemia. Blood. 2022;140(Supplement 1):1427–1429.

53. Rausch C, Rothenberg-Thurley M, Dufour AM, et al. Validation of the 2022 European Leukemianet Genetic Risk Stratification of Acute Myeloid Leukemia. Blood. 2022;140(Supplement 1):3408–3409.

54. Mecklenbrauck R, Borchert N, Funke C, et al. Prognostic Impact of Clonal Hierarchy of Myelodysplasia-Related Gene Mutations in AML Patients. Blood. 2022;140(Supplement 1):6254–6255.

55. Loghavi S, Reville PK, Lachowiez CA, et al. The Adverse Effect of Myelodysplasia-Related Mutations in De Novo Acute Myeloid Leukemia Is Associated with Higher Variant Allelic Frequency (VAF): A Proposal for a Numeric Cutoff for Variant Allelic Frequency. Blood. 2022;140(Supplement 1):6306–6308.

