## supplementary figures for "Interaction between myelodysplasia-related gene mutations and ontogeny in acute myeloid leukemia: an appraisal of the new WHO and IC classifications and ELN risk stratification"

Running title: acute myeloid leukemia with myelodysplasia related gene mutations

Authors: Joseph GW. McCarter[1,2,3], David Nemirovsky[4], Christopher A. Famulare[3], Noushin Farnoud[1,3], Abhinita S. Mohanty[5], Zoe S. Stone-Molloy[3], Jordan Chervin[3], Brian Ball[6], Zachary D. Epstein-Peterson[6], Maria E. Arcila[5], Aaron J. Stonestrom[3,6], Andrew Dunbar[3,6], Sheng F. Cai[3,6], Jacob L. Glass[3,6], Mark B. Geyer[3,6], Raajit K. Rampal[3,6], Ellin Berman[3,6], Omar I. Abdel-Wahab[3,6,7], Eytan M. Stein[3,6], Martin S. Tallman[3,6], Ross L. Levine[3,6,7], Aaron D. Goldberg[3,6], Elli Papaemmanuil[1,3], Yanming Zhang[8], Mikhail Roshal[9], Andriy Derkach[4], Wenbin Xiao[3,9]

Supplemental Figures 1-11

supplemental Figure 1

A

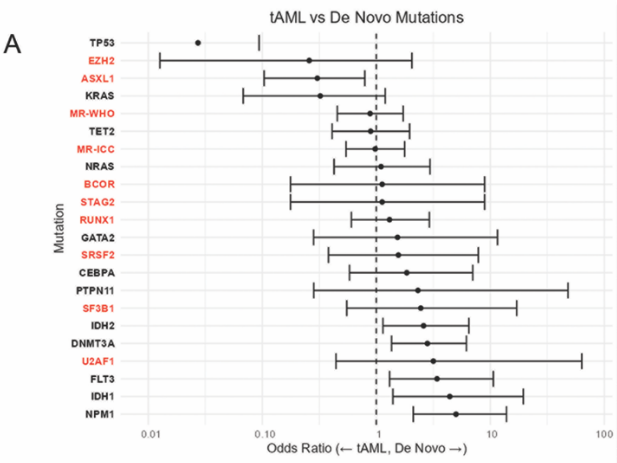


tAML vs De Novo Mutations

| Mutation | tAML, N = 92[1] | De Novo, N = 116[1] | OR[2] | 95% CI[2] | p-value[3] |
|---|---|---|---|---|---|
| TP53 | 36/92 (39%) | 2/116 (1.7%) | 0.03 | 0.00, 0.09 | **<0.001** |
| EZH2 | 3/92 (3.3%) | 1/116 (0.9%) | 0.26 | 0.01, 2.05 | 0.3 |
| ASXL1 | 14/92 (15%) | 6/116 (5.2%) | 0.3 | 0.10, 0.79 | **0.018** |
| KRAS | 7/92 (7.6%) | 3/116 (2.6%) | 0.32 | 0.07, 1.20 | 0.11 |
| MR-WHO | 21/92 (23%) | 24/116 (21%) | 0.88 | 0.45, 1.72 | 0.7 |
| TET2 | 14/92 (15%) | 16/116 (14%) | 0.89 | 0.41, 1.96 | 0.8 |
| MR-ICC | 29/92 (32%) | 36/116 (31%) | 0.98 | 0.54, 1.77 | >0.9 |
| NRAS | 8/92 (8.7%) | 11/116 (9.5%) | 1.1 | 0.43, 2.96 | >0.9 |
| BCOR | 2/35 (5.7%) | 3/47 (6.4%) | 1.13 | 0.18, 8.91 | >0.9 |
| STAG2 | 2/35 (5.7%) | 3/47 (6.4%) | 1.13 | 0.18, 8.91 | >0.9 |
| RUNX1 | 12/92 (13%) | 19/116 (16%) | 1.31 | 0.60, 2.92 | 0.6 |
| GATA2 | 2/35 (5.7%) | 4/47 (8.5%) | 1.53 | 0.28, 11.6 | >0.9 |
| SRSF2 | 3/35 (8.6%) | 6/47 (13%) | 1.56 | 0.38, 7.85 | 0.7 |
| CEBPA | 4/92 (4.3%) | 9/116 (7.8%) | 1.85 | 0.58, 7.02 | 0.4 |
| PTPN11 | 1/35 (2.9%) | 3/47 (6.4%) | 2.32 | 0.28, 48.0 | 0.6 |
| SF3B1 | 2/92 (2.2%) | 6/116 (5.2%) | 2.45 | 0.55, 17.0 | 0.3 |
| IDH2 | 8/92 (8.7%) | 23/116 (20%) | 2.6 | 1.14, 6.48 | **0.031** |
| DNMT3A | 11/92 (12%) | 32/116 (28%) | 2.81 | 1.36, 6.17 | **0.006** |
| U2AF1 | 1/35 (2.9%) | 4/47 (8.5%) | 3.16 | 0.44, 63.4 | 0.4 |
| FLT3 | 5/92 (5.4%) | 19/116 (16%) | 3.41 | 1.31, 10.6 | **0.016** |
| IDH1 | 3/92 (3.3%) | 15/116 (13%) | 4.41 | 1.40, 19.5 | **0.014** |
| NPM1 | 6/92 (6.5%) | 30/116 (26%) | 5 | 2.11, 13.9 | **<0.001** |
| ZRSR2 | 0/35 (0%) | 1/47 (2.1%) | NE[4] | NE[4] | >0.9 |

[1] n/N (%)
[2] OR = Odds Ratio, CI = Confidence Interval
[3] Fisher's exact test
[4] Not Estimable

B

MR-Hx vs De Novo Mutations

| Mutation | MR-Hx, N = 107[1] | De Novo, N = 116[1] | OR[2] | 95% CI[2] | p-value[3] |
|---|---|---|---|---|---|
| EZH2 | 8/107 (7.5%) | 1/116 (0.9%) | 0.11 | 0.01, 0.60 | **0.015** |
| TP53 | 13/107 (12%) | 2/116 (1.7%) | 0.13 | 0.02, 0.47 | **0.002** |
| KRAS | 13/107 (12%) | 3/116 (2.6%) | 0.19 | 0.04, 0.62 | **0.008** |
| ASXL1 | 21/107 (20%) | 6/116 (5.2%) | 0.22 | 0.08, 0.55 | **<0.001** |
| MR-WHO | 55/107 (51%) | 24/116 (21%) | 0.25 | 0.14, 0.44 | **<0.001** |
| SF3B1 | 19/107 (18%) | 6/116 (5.2%) | 0.25 | 0.09, 0.63 | **0.005** |
| SRSF2 | 13/38 (34%) | 6/47 (13%) | 0.28 | 0.09, 0.81 | **0.034** |
| MR-ICC | 65/107 (61%) | 36/116 (31%) | 0.29 | 0.17, 0.50 | **<0.001** |
| ZRSR2 | 2/38 (5.3%) | 1/47 (2.1%) | 0.39 | 0.02, 4.24 | 0.6 |
| RUNX1 | 32/107 (30%) | 19/116 (16%) | 0.46 | 0.24, 0.87 | **0.017** |
| TET2 | 24/107 (22%) | 16/116 (14%) | 0.55 | 0.27, 1.10 | 0.12 |
| PTPN11 | 4/38 (11%) | 3/47 (6.4%) | 0.58 | 0.11, 2.80 | 0.7 |
| STAG2 | 4/38 (11%) | 3/47 (6.4%) | 0.58 | 0.11, 2.80 | 0.7 |
| U2AF1 | 5/38 (13%) | 4/47 (8.5%) | 0.61 | 0.14, 2.49 | 0.5 |
| NRAS | 14/107 (13%) | 11/116 (9.5%) | 0.7 | 0.30, 1.60 | 0.4 |
| IDH1 | 9/107 (8.4%) | 15/116 (13%) | 1.62 | 0.69, 4.01 | 0.3 |
| FLT3 | 11/107 (10%) | 19/116 (16%) | 1.71 | 0.78, 3.90 | 0.2 |
| DNMT3A | 18/107 (17%) | 32/116 (28%) | 1.88 | 0.99, 3.66 | 0.076 |
| CEBPA | 4/107 (3.7%) | 9/116 (7.8%) | 2.17 | 0.68, 8.19 | 0.3 |
| BCOR | 1/38 (2.6%) | 3/47 (6.4%) | 2.52 | 0.31, 52.2 | 0.6 |
| GATA2 | 1/38 (2.6%) | 4/47 (8.5%) | 3.44 | 0.48, 68.9 | 0.4 |
| IDH2 | 7/107 (6.5%) | 23/116 (20%) | 3.53 | 1.52, 9.26 | **0.005** |
| NPM1 | 7/107 (6.5%) | 30/116 (26%) | 4.98 | 2.19, 12.9 | **<0.001** |

[1] n/N (%)
[2] OR = Odds Ratio, CI = Confidence Interval
[3] Fisher's exact test

C

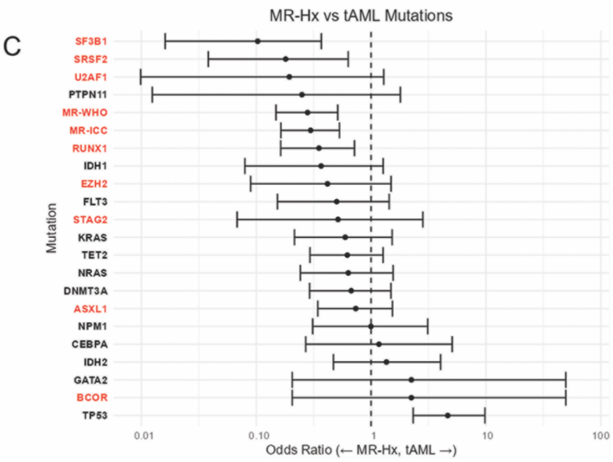


MR-Hx vs tAML Mutations

| Mutation | MR-Hx, N = 107[1] | tAML, N = 92[1] | OR[2] | 95% CI[2] | p-value[3] |
|---|---|---|---|---|---|
| ZRSR2 | 2/38 (5.3%) | 0/35 (0%) | NE[4] | NE[4] | 0.5 |
| SF3B1 | 19/107 (18%) | 2/92 (2.2%) | 0.1 | 0.02, 0.37 | **<0.001** |
| SRSF2 | 13/38 (34%) | 3/35 (8.6%) | 0.18 | 0.04, 0.63 | **0.011** |
| U2AF1 | 5/38 (13%) | 1/35 (2.9%) | 0.19 | 0.01, 1.29 | 0.2 |
| PTPN11 | 4/38 (11%) | 1/35 (2.9%) | 0.25 | 0.01, 1.80 | 0.4 |
| MR-WHO | 55/107 (51%) | 21/92 (23%) | 0.28 | 0.15, 0.51 | **<0.001** |
| MR-ICC | 65/107 (61%) | 29/92 (32%) | 0.3 | 0.16, 0.53 | **<0.001** |
| RUNX1 | 32/107 (30%) | 12/92 (13%) | 0.35 | 0.16, 0.72 | **0.006** |
| IDH1 | 9/107 (8.4%) | 3/92 (3.3%) | 0.37 | 0.08, 1.27 | 0.15 |
| EZH2 | 8/107 (7.5%) | 3/92 (3.3%) | 0.42 | 0.09, 1.49 | 0.2 |
| FLT3 | 11/107 (10%) | 5/92 (5.4%) | 0.5 | 0.15, 1.44 | 0.3 |
| STAG2 | 4/38 (11%) | 2/35 (5.7%) | 0.52 | 0.07, 2.83 | 0.7 |
| KRAS | 13/107 (12%) | 7/92 (7.6%) | 0.6 | 0.21, 1.52 | 0.3 |
| TET2 | 24/107 (22%) | 14/92 (15%) | 0.62 | 0.29, 1.27 | 0.2 |
| NRAS | 14/107 (13%) | 8/92 (8.7%) | 0.63 | 0.24, 1.55 | 0.4 |
| DNMT3A | 18/107 (17%) | 11/92 (12%) | 0.67 | 0.29, 1.49 | 0.4 |
| ASXL1 | 21/107 (20%) | 14/92 (15%) | 0.74 | 0.34, 1.53 | 0.5 |
| NPM1 | 7/107 (6.5%) | 6/92 (6.5%) | 1 | 0.31, 3.11 | >0.9 |
| CEBPA | 4/107 (3.7%) | 4/92 (4.3%) | 1.17 | 0.27, 5.08 | >0.9 |
| IDH2 | 7/107 (6.5%) | 8/92 (8.7%) | 1.36 | 0.47, 4.03 | 0.6 |
| GATA2 | 1/38 (2.6%) | 2/35 (5.7%) | 2.24 | 0.21, 49.6 | 0.6 |
| BCOR | 1/38 (2.6%) | 2/35 (5.7%) | 2.24 | 0.21, 49.6 | 0.6 |
| TP53 | 13/107 (12%) | 36/92 (39%) | 4.65 | 2.32, 9.80 | **<0.001** |
| NOTCH2 | 0/10 (0%) | 2/17 (12%) | NE[4] | NE[4] | 0.5 |

[1] n/N (%)
[2] OR = Odds Ratio, CI = Confidence Interval
[3] Fisher's exact test
[4] Not Estimable

Supplemental Figure 1. Association between individual gene mutations and AML subtypes defined by ontogeny, as depicted by odds ratio on a $log_{10}$ scale. Both forest plots and tables are shown. A. t-AML vs *de novo* AML. B. MR-Hx vs *de novo* AML (also see Figure 1C). C. MR-Hx vs t-AML. *MR/RUNX1* genes are highlighted in red.

Supplemental Figure 2

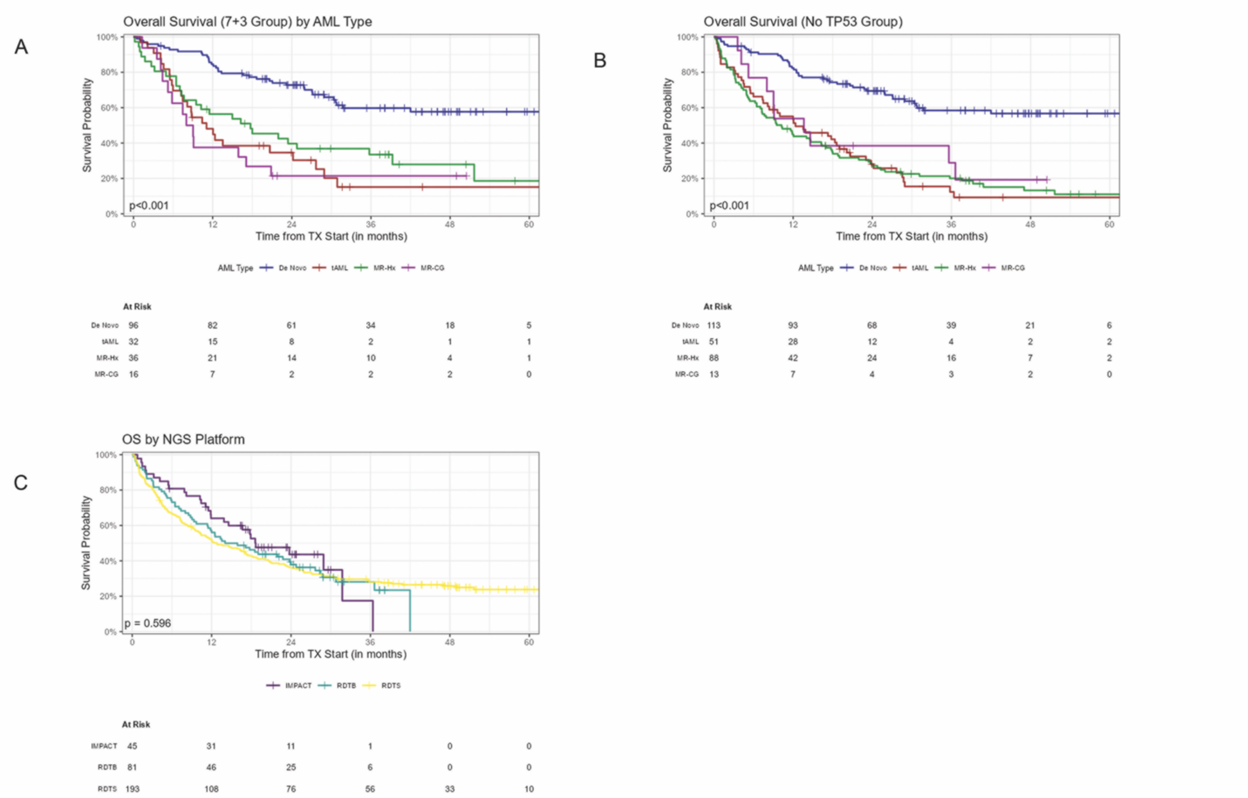


Supplemental Figure 2. A-B. Kaplan-Meier curves showing overall survival of AML patients divided by ontogeny. A. AML patients treated with 7+3 induction regimen. B. AML patients with non-*TP53* mutations (*TP53* mutated AML excluded). C. Kaplan-Meier curves of overall survival of AML patients sequenced by different NGS platforms.

supplemental FIgure 3

A

Cox PH model adjusting for age (as a cubic spline), AML type, treatment, transplant, and TP53 mutation; stratified by ELN 2022 risk

| Characteristic | HR[1] | 95% CI[1] | p-value |
|---|---|---|---|
| **AML Type** | | | **0.003** |
| De Novo | Ref. | Ref. | |
| tAML | 1.93 | 1.24, 2.99 | |
| MR-Hx | 2.15 | 1.43, 3.24 | |
| MR-CG | 1.93 | 1.11, 3.34 | |
| **Age[2]** | | | **<0.001** |
| **Treatment** | | | **<0.001** |
| 7+3 | Ref. | Ref. | |
| Venetoclax + HMA/low DAC | 0.87 | 0.47, 1.62 | |
| HMA | 0.82 | 0.52, 1.31 | |
| Trial | 1.15 | 0.75, 1.77 | |
| None/Supportive Care | 6.00 | 3.46, 10.4 | |
| **Transplant** | 0.58 | 0.39, 0.88 | **0.009** |
| **TP53** | 1.62 | 1.15, 2.28 | **0.006** |

[1] HR = Hazard Ratio, CI = Confidence Interval
[2] A cubic spline was used for the age covariate

B

Cox PH model adjusting for age (as a cubic spline), AML type, treatment, transplant, and TP53 mutation; stratified by genomic class

| Characteristic | HR[1] | 95% CI[1] | p-value |
|---|---|---|---|
| **AML Type** | | | **0.001** |
| De Novo | Ref. | Ref. | |
| tAML | 2.11 | 1.33, 3.36 | |
| MR-Hx | 2.35 | 1.52, 3.63 | |
| MR-CG | 2.30 | 1.30, 4.06 | |
| **Age[2]** | | | **<0.001** |
| **Treatment** | | | **<0.001** |
| 7+3 | Ref. | Ref. | |
| Venetoclax + HMA/low DAC | 0.84 | 0.44, 1.59 | |
| HMA | 0.87 | 0.54, 1.38 | |
| Trial | 1.15 | 0.74, 1.79 | |
| None/Supportive Care | 6.18 | 3.48, 11.0 | |
| **Transplant** | 0.62 | 0.41, 0.94 | **0.023** |
| **TP53** | 1.29 | 0.31, 5.38 | 0.7 |

[1] HR = Hazard Ratio, CI = Confidence Interval
[2] A cubic spline was used for the age covariate

C

Cox PH model adjusting for age (as a cubic spline), AML type, treatment, transplant, and TP53 mutation; stratified by cytogenetic risk

| Characteristic | HR[1] | 95% CI[1] | p-value |
|---|---|---|---|
| **AML Type** | | | **0.004** |
| De Novo | Ref. | Ref. | |
| tAML | 2.05 | 1.30, 3.24 | |
| MR-Hx | 2.20 | 1.42, 3.40 | |
| MR-CG | 1.89 | 1.03, 3.49 | |
| **Age[2]** | | | **<0.001** |
| **Treatment** | | | **<0.001** |
| 7+3 | Ref. | Ref. | |
| Venetoclax + HMA/low DAC | 0.84 | 0.45, 1.57 | |
| HMA | 0.83 | 0.52, 1.32 | |
| Trial | 1.21 | 0.79, 1.87 | |
| None/Supportive Care | 6.32 | 3.63, 11.0 | |
| **Transplant** | 0.59 | 0.40, 0.88 | **0.010** |
| **TP53** | 1.44 | 0.98, 2.12 | 0.065 |

[1] HR = Hazard Ratio, CI = Confidence Interval
[2] A cubic spline was used for the age covariate

D

Cox PH model adjusting for age (as a cubic spline), genomic class, treatment, and transplant; stratified by AML type

| Characteristic | HR[1] | 95% CI[1] | p-value |
|---|---|---|---|
| **Molecular Group** | | | **0.003** |
| inv16, t(15;17), t(8;21) | Ref. | Ref. | |
| CEBPA | 1.03 | 0.18, 5.90 | |
| NPM1 | 4.21 | 1.36, 13.0 | |
| MLL | 3.95 | 1.04, 15.0 | |
| t(6;9) | NE[2] | NE[2] | |
| EVI1 | 6.53 | 2.02, 21.1 | |
| TP53 | 6.73 | 2.28, 19.9 | |
| MR or RUNX1 | 4.11 | 1.39, 12.2 | |
| NOS | 3.34 | 1.12, 10.0 | |
| NEG | 3.19 | 0.90, 11.3 | |
| **Age[3]** | | | **<0.001** |
| **Treatment** | | | **<0.001** |
| 7+3 | Ref. | Ref. | |
| Venetoclax + HMA/low DAC | 0.83 | 0.44, 1.56 | |
| HMA | 0.87 | 0.55, 1.39 | |
| Trial | 1.22 | 0.79, 1.89 | |
| None/Supportive Care | 5.75 | 3.26, 10.1 | |
| **Transplant** | 0.61 | 0.41, 0.92 | **0.018** |

[1] HR = Hazard Ratio, CI = Confidence Interval
[2] Not Estimable; No t(6;9) patients experienced an event
[3] A cubic spline was used for the age covariate

Supplemental Figure 3. Multivariate analysis of AML ontogeny associated risks in AML. AML ontogeny related risk was evaluated in multivariable Cox-regression models adjusting for age (modeled by cubic spline), initial treatment at diagnosis, allogenic transplant (modeled as time-dependent variable), presence of *TP53* mutation and then stratified by ELN2022 risk (A), genomic classes (B), and cytogenetic risks (C). D. Genomic classes related risk was evaluated in a similar way and stratified by AML ontogeny. Abbreviations: 7+3, daunorubicin+cytarabine including CPX-351; HMA, hypomethylating agents; Low DAC, low-dose cytarabine.

supplemental Figure 4

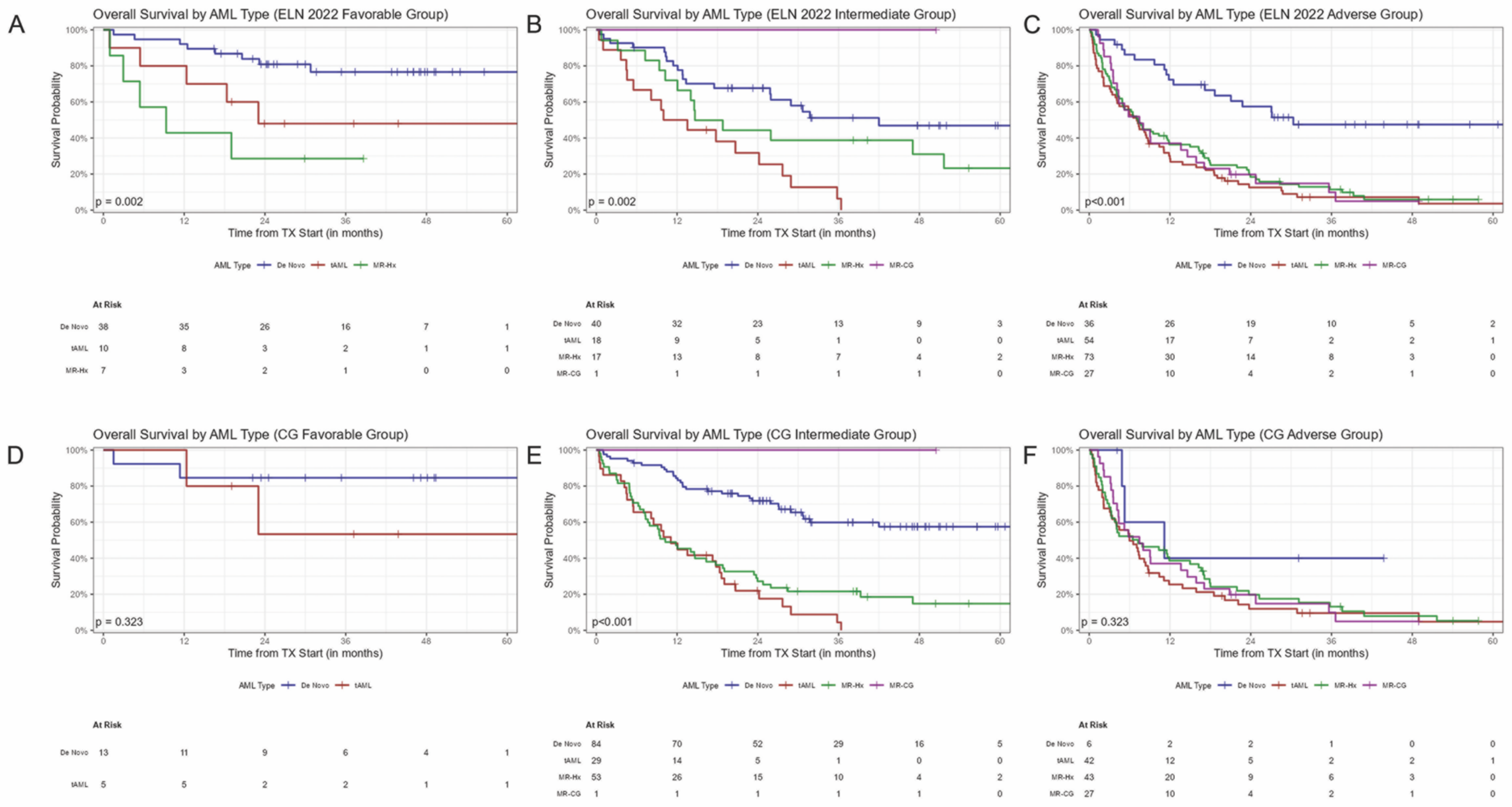


Supplemental Figure 4. Impacts of AML ontogeny on overall survival in ELN2022 and CG risk groups. A-C. Kaplan-Meier curves of overall survival of AML patients divided by ELN2022 risks and ontogeny. D-F. Kaplan-Meier curves of overall survival of AML patients divided by CG risks and ontogeny.

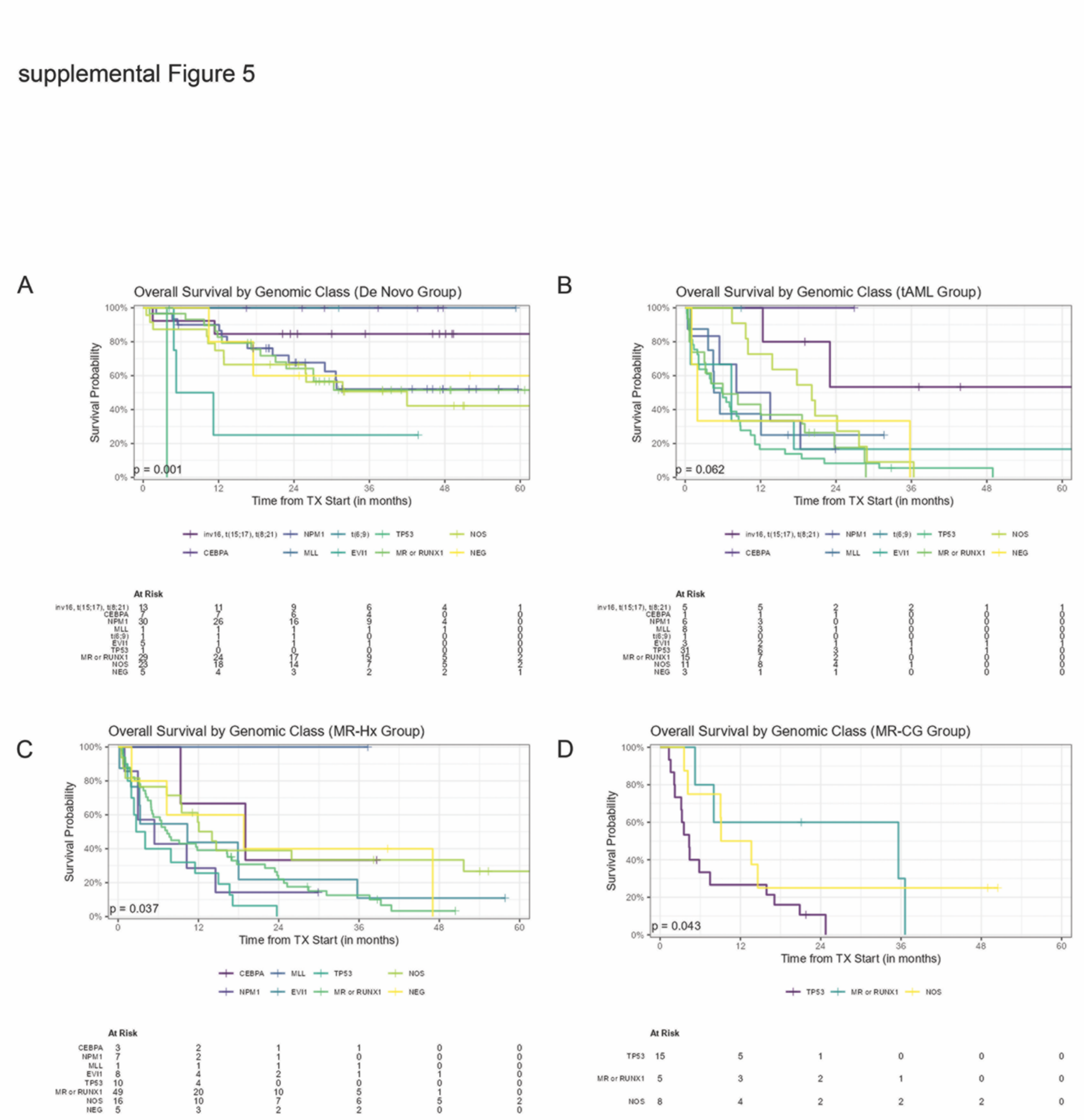


Supplemental Figure 5. Overall survival of AML patients divided by genomic classes. A, *de novo* AML subgroup. B, t-AML subgroup. C, MR-Hx AML subgroup. D, MR-CG AML subgroup.

supplemental Figure 6

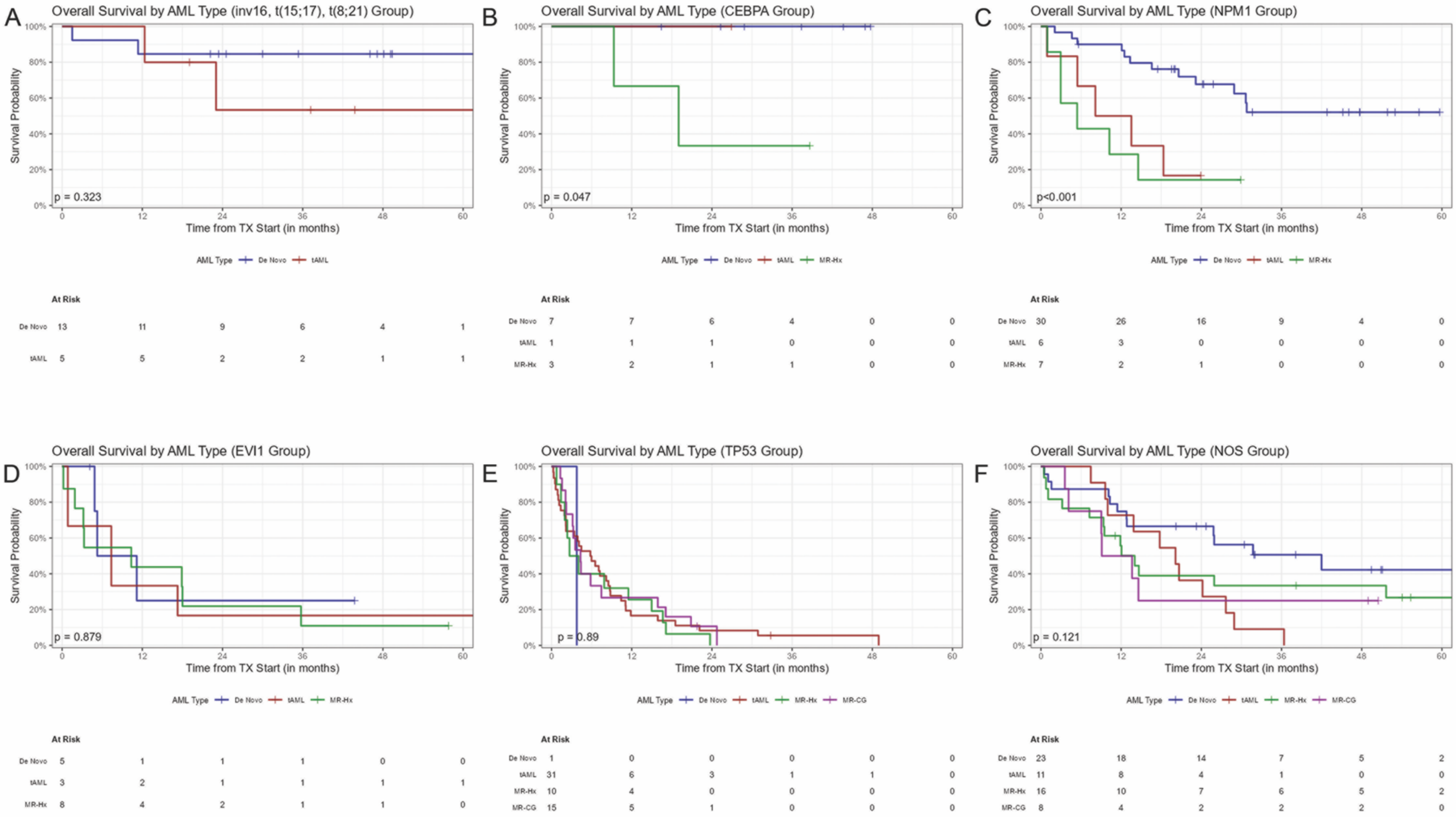


Supplemental Figure 6. Impact of AML ontogeny on overall survival of AML patients in each genomic class. A, AML with favorable fusions. B, *CEBPA bZIP* mutated AML. C, *NPM1* mutated AML. D, *EVI1* rearranged AML. E, *TP53* mutated AML. F, AML with non-class defining mutations/rearrangements (NOS).

supplemental Figure 7

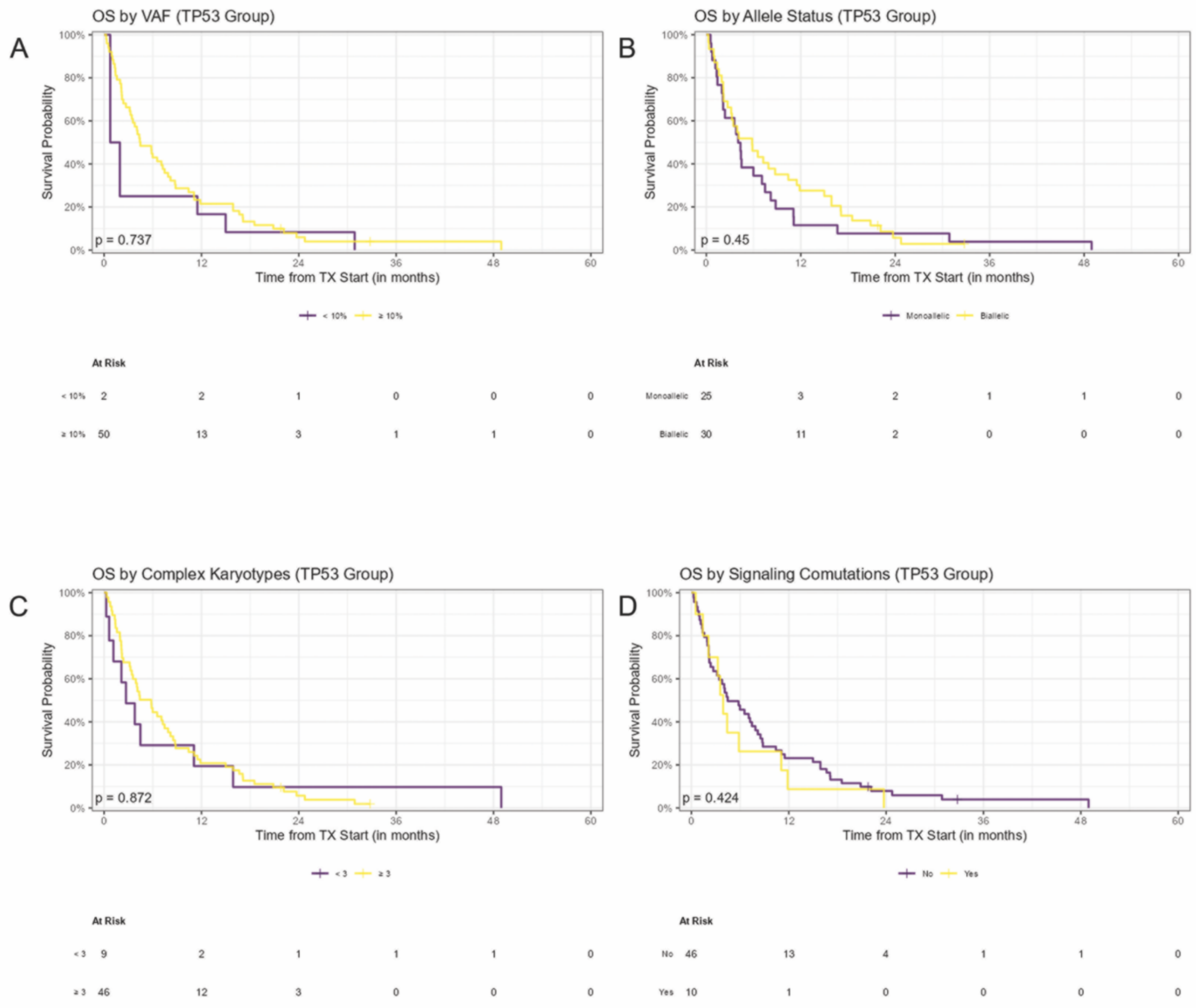


Supplemental Figure 7. Impact of *TP53* mutation burden (A, variant allelic frequency <10% vs >=10%), allelic status (B, mono- vs bi- or multi hits), complex karyotype (C) and co-mutations in signaling molecules (D). Bi- or multi-hits of *TP53* are defined as more than one mutations, or one or more mutations with del17p/-17. Co-mutations in signaling molecules include mutations in *NRAS, KRAS, CBL, BRAF, FLT3 and PTPN11*.

supplemental FIgure 8

A

**Univariate OS by Mutation**

| Characteristic | Mutation Count[1] | HR[2] | 95% CI[2] | p-value |
|---|---|---|---|---|
| DNMT3A | 67/344 (19%) | 0.70 | 0.50, 0.98 | **0.040** |
| TP53 | 67/344 (19%) | 2.84 | 2.12, 3.79 | **<0.001** |
| TET2 | 57/344 (17%) | 1.28 | 0.92, 1.77 | 0.14 |
| NPM1 | 43/344 (12%) | 0.61 | 0.40, 0.94 | **0.025** |
| IDH2 | 40/344 (12%) | 0.65 | 0.42, 0.99 | **0.043** |
| NRAS | 38/344 (11%) | 0.98 | 0.66, 1.46 | >0.9 |
| FLT3 | 36/344 (10%) | 0.89 | 0.58, 1.36 | 0.6 |
| IDH1 | 28/344 (8.1%) | 0.87 | 0.55, 1.39 | 0.6 |
| KRAS | 23/344 (6.7%) | 2.11 | 1.32, 3.39 | **0.002** |
| PTPN11 | 8/132 (6.1%) | 1.25 | 0.58, 2.72 | 0.6 |
| NOTCH2 | 3/50 (6.0%) | 3.91 | 1.13, 13.5 | **0.032** |
| GATA2 | 7/132 (5.3%) | 0.43 | 0.13, 1.35 | 0.15 |
| CEBPA | 18/344 (5.2%) | 0.36 | 0.17, 0.77 | **0.008** |
| **MR-WHO** | 107/344 (31%) | 1.48 | 1.13, 1.92 | **0.004** |
| ASXL1 | 45/344 (13%) | 1.23 | 0.86, 1.76 | 0.3 |
| BCOR | 7/132 (5.3%) | 1.61 | 0.65, 3.99 | 0.3 |
| EZH2 | 12/344 (3.5%) | 1.92 | 1.05, 3.53 | **0.035** |
| STAG2 | 10/132 (7.6%) | 1.27 | 0.61, 2.64 | 0.5 |
| SF3B1 | 28/344 (8.1%) | 1.71 | 1.14, 2.58 | **0.010** |
| SRSF2 | 23/132 (17%) | 1.24 | 0.73, 2.13 | 0.4 |
| ZRSR2 | 3/132 (2.3%) | 1.28 | 0.31, 5.22 | 0.7 |
| U2AF1 | 11/132 (8.3%) | 1.85 | 0.92, 3.72 | 0.086 |
| **MR-ICC** | 138/344 (40%) | 1.33 | 1.03, 1.72 | **0.029** |
| RUNX1 | 65/344 (19%) | 1.16 | 0.85, 1.59 | 0.4 |

[1] n/N (%)
[2] HR = Hazard Ratio, CI = Confidence Interval

B

**OS by Mutations, Adjusted for AML Type**

| Characteristic | Mutation Count[1] | HR[2] | 95% CI[2] | p-value |
|---|---|---|---|---|
| DNMT3A | 67/344 (19%) | 0.86 | 0.61, 1.22 | 0.4 |
| TP53 | 67/344 (19%) | 1.93 | 1.39, 2.69 | **<0.001** |
| TET2 | 57/344 (17%) | 1.14 | 0.82, 1.58 | 0.4 |
| NPM1 | 43/344 (12%) | 1.12 | 0.72, 1.77 | 0.6 |
| IDH2 | 40/344 (12%) | 0.78 | 0.51, 1.20 | 0.3 |
| NRAS | 38/344 (11%) | 0.88 | 0.59, 1.32 | 0.5 |
| FLT3 | 36/344 (10%) | 1.23 | 0.80, 1.90 | 0.3 |
| IDH1 | 28/344 (8.1%) | 1.07 | 0.66, 1.72 | 0.8 |
| KRAS | 23/344 (6.7%) | 1.55 | 0.96, 2.51 | 0.076 |
| PTPN11 | 8/132 (6.1%) | 1.21 | 0.55, 2.68 | 0.6 |
| NOTCH2 | 3/50 (6.0%) | 2.14 | 0.54, 8.54 | 0.3 |
| GATA2 | 7/132 (5.3%) | 0.50 | 0.16, 1.62 | 0.2 |
| CEBPA | 18/344 (5.2%) | 0.42 | 0.20, 0.90 | **0.025** |
| **MR-WHO** | 107/344 (31%) | 1.19 | 0.89, 1.58 | 0.2 |
| ASXL1 | 45/344 (13%) | 0.90 | 0.63, 1.31 | 0.6 |
| BCOR | 7/132 (5.3%) | 2.12 | 0.84, 5.36 | 0.11 |
| EZH2 | 12/344 (3.5%) | 1.43 | 0.77, 2.66 | 0.3 |
| STAG2 | 10/132 (7.6%) | 1.30 | 0.59, 2.88 | 0.5 |
| SF3B1 | 28/344 (8.1%) | 1.42 | 0.91, 2.19 | 0.12 |
| SRSF2 | 23/132 (17%) | 0.94 | 0.53, 1.69 | 0.8 |
| ZRSR2 | 3/132 (2.3%) | 1.61 | 0.38, 6.81 | 0.5 |
| U2AF1 | 11/132 (8.3%) | 1.80 | 0.87, 3.73 | 0.11 |
| **MR-ICC** | 138/344 (40%) | 1.16 | 0.89, 1.52 | 0.3 |
| RUNX1 | 65/344 (19%) | 1.13 | 0.82, 1.56 | 0.5 |

[1] n/N (%)
[2] HR = Hazard Ratio, CI = Confidence Interval

C

**Cox PH model adjusting for age (as a cubic spline), treatment, transplant, AML type, and ELN 2022 risk**

| Characteristic | HR[1] | 95% CI[1] | p-value |
|---|---|---|---|
| **Age**[2] | | | **<0.001** |
| **Treatment** | | | **<0.001** |
| 7+3 | Ref. | Ref. | |
| Venetoclax + HMA/low DAC | 0.82 | 0.44, 1.52 | |
| HMA | 0.86 | 0.54, 1.36 | |
| Trial | 1.25 | 0.82, 1.92 | |
| None/Supportive Care | 6.27 | 3.64, 10.8 | |
| **Transplant** | 0.60 | 0.40, 0.90 | **0.012** |
| **AML Type** | | | **<0.001** |
| De Novo | Ref. | Ref. | |
| tAML | 2.31 | 1.53, 3.49 | |
| MR-Hx | 2.24 | 1.50, 3.35 | |
| MR-CG | 2.16 | 1.26, 3.70 | |
| **ELN Risk** | | | **<0.001** |
| Favorable | Ref. | Ref. | |
| Intermediate | 1.71 | 0.97, 3.01 | |
| Adverse | 2.57 | 1.50, 4.38 | |

[1] HR = Hazard Ratio, CI = Confidence Interval
[2] A cubic spline was used for the age covariate

Supplemental Figure 8. A-B, Univariable Cox proportional hazard regression to evaluate OS associated with individual mutations with (B) or without (A) adjusting for AML ontogeny. C, Multivariate analysis on overall survival of the entire cohort of AML patients adjusting for age, treatment, transplant (allo-HSCT), AML ontogeny, and ELN risks. Abbreviations: 7+3, daunorubicin+cytarabine including CPX-351; HMA, hypomethylating agents; Low DAC, low-dose cytarabine.

supplemental Figure 9

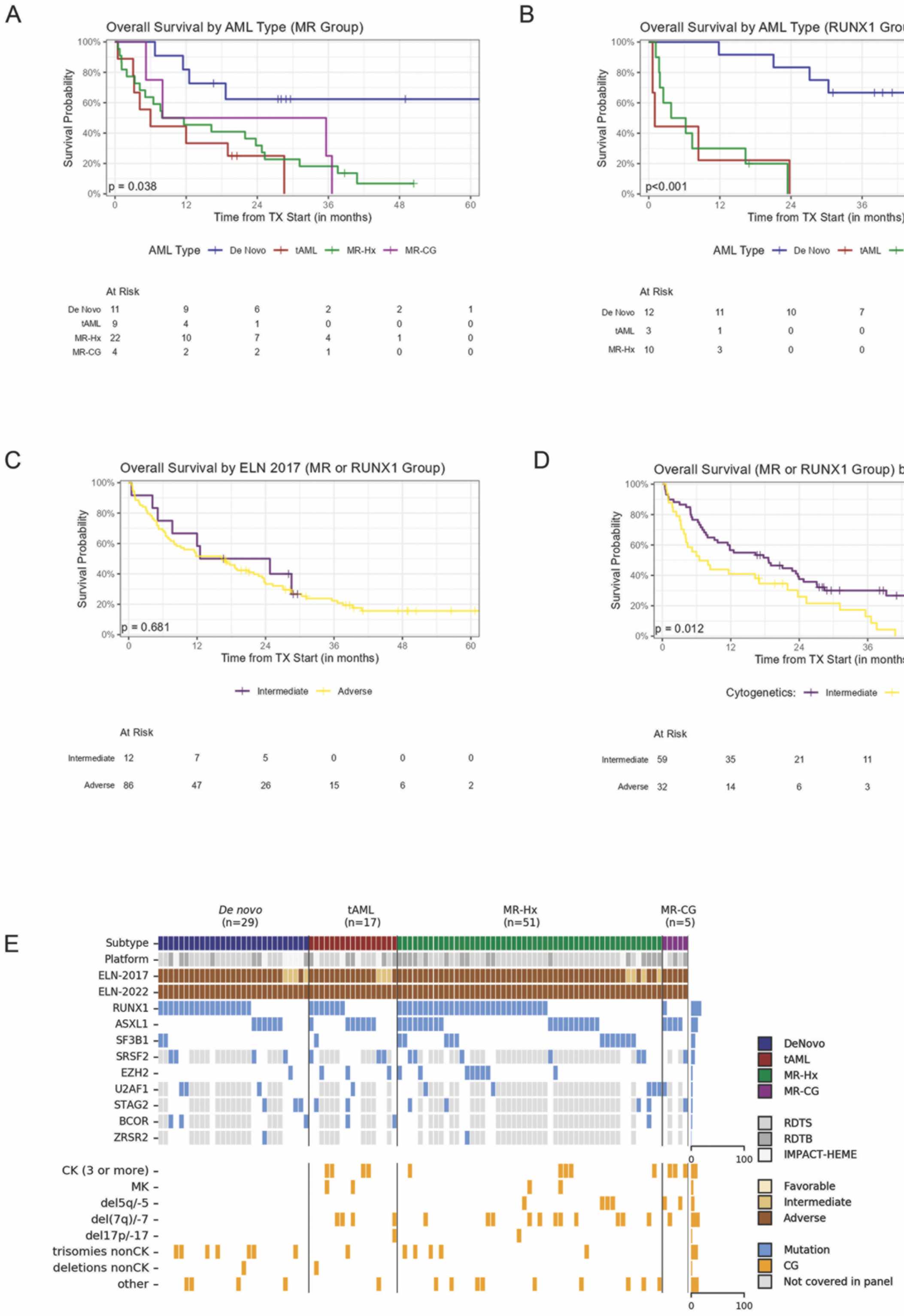

Supplemental Figure 9. A, Kaplan-Meier curves of overall survival of AML with MR (no *RUNX1*) gene mutations divided by ontogeny. B, A. Kaplan-Meier curves of overall survival of AML with *RUNX1* (no other MR) gene mutations divided by ontogeny. C-D, A. Kaplan-Meier curves of overall survival of AML with *MR/RUNX1* gene mutations divided by ELN2017 risks (C) and CG risks (D). E, Oncoplot of AML with *MR/RUNX1* mutations.

supplemental Figure 10

A

OS by Mutations, Adjusted for AML Type

| Characteristic | Mutation Count[1] | HR[2] | 95% CI[2] | p-value |
|---|---|---|---|---|
| DNMT3A | 21/102 (21%) | 0.84 | 0.46, 1.54 | 0.6 |
| TP53 | 0/102 (0%) | | | |
| TET2 | 22/102 (22%) | 1.36 | 0.81, 2.29 | 0.2 |
| NPM1 | 0/102 (0%) | | | |
| IDH2 | 15/102 (15%) | 0.51 | 0.24, 1.08 | 0.078 |
| NRAS | 13/102 (13%) | 3.44 | 1.70, 6.94 | **<0.001** |
| FLT3 | 14/102 (14%) | 1.76 | 0.95, 3.26 | 0.072 |
| IDH1 | 5/102 (4.9%) | 0.89 | 0.35, 2.25 | 0.8 |
| KRAS | 9/102 (8.8%) | 2.72 | 1.31, 5.67 | **0.008** |
| PTPN11 | 5/43 (12%) | 1.19 | 0.44, 3.19 | 0.7 |
| NOTCH2 | 0/13 (0%) | | | |
| GATA2 | 2/43 (4.7%) | 0.16 | 0.02, 1.43 | 0.10 |
| CEBPA | 3/102 (2.9%) | 0.70 | 0.15, 3.20 | 0.6 |
| **MR-WHO** | 76/102 (75%) | 0.83 | 0.46, 1.51 | 0.5 |
| ASXL1 | 36/102 (35%) | 0.63 | 0.39, 1.03 | 0.068 |
| BCOR | 6/43 (14%) | 1.50 | 0.49, 4.58 | 0.5 |
| EZH2 | 12/102 (12%) | 1.37 | 0.71, 2.64 | 0.3 |
| STAG2 | 10/43 (23%) | 1.44 | 0.59, 3.53 | 0.4 |
| SF3B1 | 15/102 (15%) | 0.94 | 0.52, 1.72 | 0.8 |
| SRSF2 | 14/43 (33%) | 1.07 | 0.49, 2.32 | 0.9 |
| ZRSR2 | 2/43 (4.7%) | 0.70 | 0.09, 5.29 | 0.7 |
| U2AF1 | 10/43 (23%) | 2.38 | 0.98, 5.82 | 0.057 |
| **MR-ICC** | 102/102 (100%) | | | |
| RUNX1 | 55/102 (54%) | 1.29 | 0.81, 2.05 | 0.3 |

[1] n/N (%)
[2] HR = Hazard Ratio, CI = Confidence Interval

B

Cox PH model adjusting for age (as a cubic spline), AML type, treatment, transplant, NRAS mutation, KRAS mutation, and cytogenetic risk

| Characteristic | HR[1] | 95% CI[1] | p-value |
|---|---|---|---|
| **AML Type** | | | **<0.001** |
| De Novo | Ref. | Ref. | |
| tAML | 10.6 | 3.41, 33.1 | |
| MR-Hx | 6.04 | 2.53, 14.4 | |
| MR-CG | 1.23 | 0.26, 5.77 | |
| **Age**[2] | | | 0.2 |
| **Treatment** | | | **<0.001** |
| 7+3 | Ref. | Ref. | |
| Venetoclax + HMA/low DAC | 0.52 | 0.15, 1.82 | |
| HMA | 0.27 | 0.10, 0.78 | |
| Trial | 1.39 | 0.54, 3.57 | |
| None/Supportive Care | 10.0 | 2.93, 34.3 | |
| **Transplant** | 0.31 | 0.13, 0.74 | **0.007** |
| **NRAS** | 2.18 | 0.90, 5.28 | 0.10 |
| **KRAS** | 1.37 | 0.55, 3.40 | 0.5 |
| **CG Risk** | | | 0.5 |
| Intermediate | Ref. | Ref. | |
| Adverse | 1.29 | 0.66, 2.54 | |

[1] HR = Hazard Ratio, CI = Confidence Interval
[2] A cubic spline was used for the age covariate

Supplemental Figure 10. A, Univariable Cox proportional hazard regression to evaluate OS associated with individual mutations adjusting for AML ontogeny in AML patients with *MR/RUNX1* mutations. B, Multivariate analysis on overall survival of AML patients with *MR/RUNX1* mutations adjusting for age, treatment, transplant (allo-HSCT), AML ontogeny, and *NRAS, KRAS* and CG risks. Abbreviations: 7+3, daunorubicin+cytarabine including CPX-351; HMA, hypomethylating agents; Low DAC, low-dose cytarabine.

supplemental Figure 11

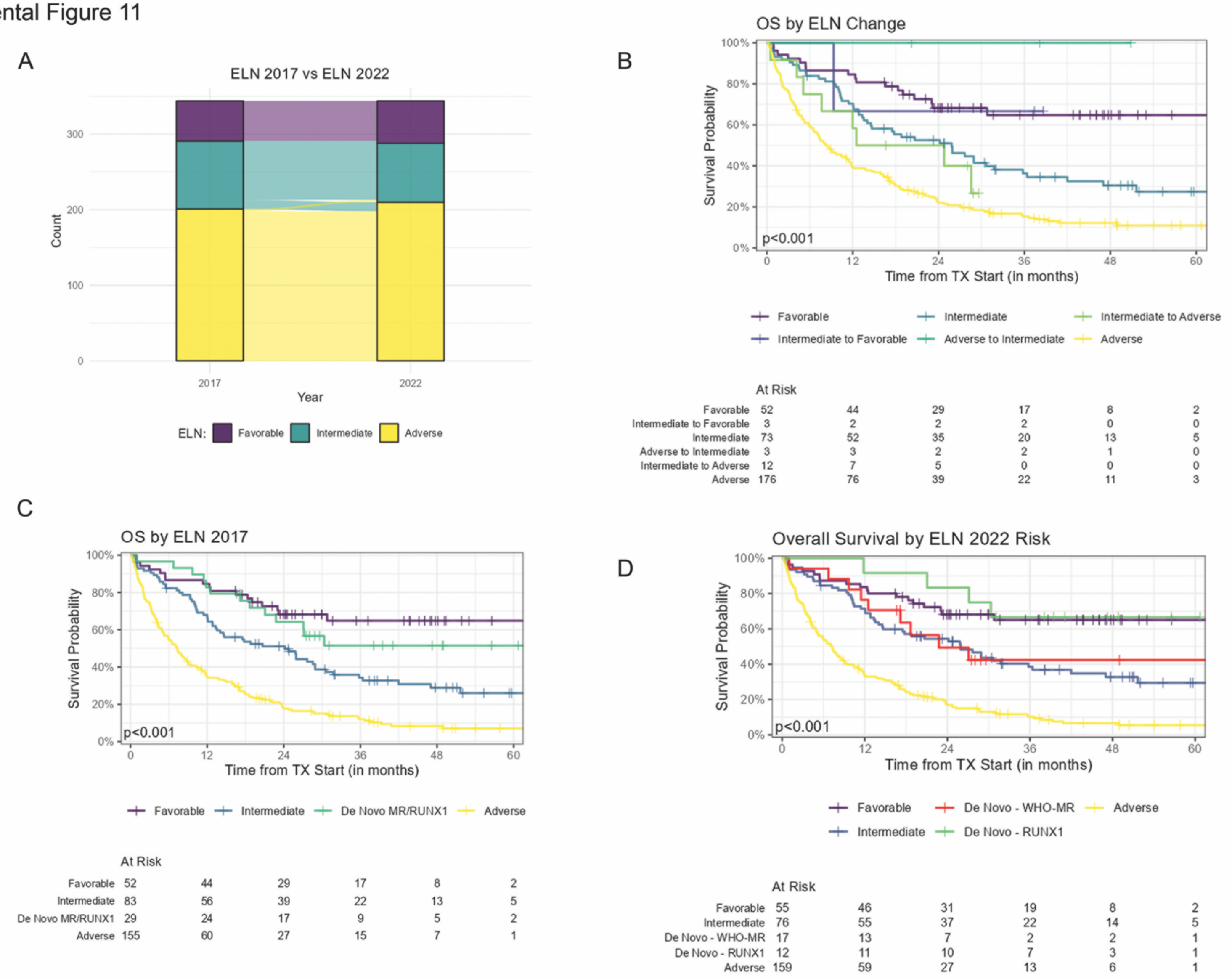

Supplemental Figure 11. A, Correlation between ELN2017 and ELN2022 risk groups. B, Overall survival of shifting risk patients between ELN2017 and ELN2022. C, *De novo* AML with *MR/RUNX1* mutations do not show adverse outcome. D. *De novo* AML with MR gene mutations show an outcome overlapping with intermediate-risk group and *de novo* AML with RUNX1 mutations only show an outcome overlapping with favorable-risk group.
